# Secreted GPNMB enhances uptake of fibrillar alpha-synuclein in a non-cell-autonomous process that can be blocked by anti-GPNMB antibodies

**DOI:** 10.64898/2026.01.26.26344888

**Authors:** Marc Carceles-Cordon, Eliza M. Brody, Masen L. Boucher, Michael D. Gallagher, Robert T. Skrinak, Travis L. Unger, Cooper K. Penner, Adama J. Berndt, Sromona Das, Katie Lam, Rudolf Jaenisch, Vivianna Van Deerlin, Edward B. Lee, Kurt Brunden, Kelvin Luk, Alice S. Chen-Plotkin

## Abstract

Glycoprotein nonmetastatic melanoma B (GPNMB), encoded by the target gene (*GPNMB*) of a Parkinson’s disease (PD) risk locus, acts as a secreted factor mediating inflammatory effects in the context of immunity and cancer. In a neurodegenerative disease context, GPNMB is critical to cellular uptake of pathological forms of alpha-synuclein (aSyn), the hallmark disease protein that misfolds and accumulates in PD. Here, we demonstrate that the non-membrane-anchored, extracellular domain of GPNMB, shed into conditioned medium or added as recombinant protein, enables uptake of aSyn fibrils in a non-cell-autonomous manner. In human postmortem brain, GPNMB is widely expressed in neurons and microglia, with increased microglial expression in the setting of neurodegenerative disease. In microglial cell lines and induced pluripotent stem cell-derived microglia (iMicroglia), GPNMB expression and secretion increases with exposure to apoptotic neurons. In the aSyn-fibril seeded model of PD, iMicroglia-derived GPNMB allows for development of aSyn pathology in *GPNMB* knockout neurons, while conditioned medium from *GPNMB* knockout iMicroglia lacks this effect. Conversely, treatment with anti-GPNMB antibodies rescues neurons from development of aSyn pathology in this model. Finally, in 1675 human postmortem cases, *GPNMB* genotypes conferring higher *GPNMB* expression associate with more widespread aSyn pathology, without affecting beta-amyloid or tau pathology. Taken together, our data suggest a positive feedback model, where neuronal death triggers increased *GPNMB* expression and secretion by microglia, leading to increased uptake of pathological forms of aSyn by neurons, leading to more neuronal death. Importantly, this cycle can be interrupted by anti-GPNMB antibodies, offering an avenue for therapeutic development.

**One Sentence Summary:** The extracellular domain of GPNMB enhances uptake of fibrillar alpha-synuclein in a non-cell-autonomous process that can be blocked by anti-GPNMB antibodies.

**Highlights:**

- The extracellular domain of GPNMB confers capacity for uptake of alpha-synuclein fibrils to iPSC-derived neurons (iNeurons) lacking *GPNMB* expression.
- GPNMB is widely expressed in neurons and microglia in human brain, with more expression in microglia, particularly in Parkinson’s disease brain.
- iPSC-derived microglia (iMicroglia) secrete GPNMB in response to neurodegeneration-related insults, and iMicroglia-derived GPNMB enhances development of alpha-synuclein pathology in iNeurons.
- Anti-GPNMB antibodies rescue iNeurons from development of synuclein pathology.
- Expression quantitative trait loci (eQTLs) for *GPNMB* associate with extent of alpha-synuclein pathology in human neurodegenerative disease.

## INTRODUCTION

Parkinson’s disease (PD), the second most common neurodegenerative disorder worldwide^1,2^, remains incurable in part due to a lack of understanding regarding the molecular events that orchestrate alpha-synuclein (aSyn) spread throughout the brain^3^. In 2022, we reported that *GPNMB*, encoding glycoprotein nonmetastatic melanoma B (GPNMB), is the target gene of a chromosome 7 PD risk locus found by genomewide association study (GWAS), with PD risk alleles associated with higher *GPNMB* expression^4^. Moreover, lowering *GPNMB* expression rescued neurons from cellular uptake of pathological forms of aSyn and subsequent development of aSyn aggregates, suggesting that GPNMB is key to the spread of aSyn pathology^4^.

Prior to its connection to PD, GPNMB was initially identified as a factor whose expression correlated with the metastatic potential of melanoma cell lines^5^. GPNMB was further studied in macrophages^6,7^, tumor cells^8^, osteoclasts^9^, and dendritic cells^10^. In these non-central nervous system (CNS) contexts, GPNMB is a Type I transmembrane glycoprotein that can span the plasma membrane^11–14^; the transmembrane form can be cleaved by metalloproteinases belonging to the ADAM10 family, to release an extracellular, non-membrane-tethered form^15–17^. This GPNMB extracellular domain (ECD) is reported to promote stemness characteristics in tumor cells^18^.

In the CNS, GPNMB has been less extensively studied, although increased expression has been reported in the context of microglial activation, especially in neurodegeneration^19–21^ and neuroinflammation^22^. In both PD^23^ and Alzheimer’s disease (AD)^24,25^, for example, *GPNMB* expression has been reported to increase as part of the disease-associated microglia (DAM) response, a transcriptomic signature first reported in a single-cell RNA profiling study of microglia found in a mouse model of AD, with transcriptionally-similar cells found in human AD^26,27^.

Here, we characterize the expression of GPNMB protein in human brain and investigate the effect of the GPNMB ECD on neuronal responses to aSyn fibrils. We present evidence for GPNMB’s role in a pathological feedback loop between degenerating neurons and microglia. Finally, we examine whether anti-GPNMB antibodies raised against the GPNMB ECD can interrupt this cycle, offering a potential therapeutic approach to disease modification in PD.

## RESULTS

### The extracellular domain of GPNMB enables uptake of aSyn fibrils in human neurons

We previously reported that GPNMB interacts with aSyn and that loss of GPNMB in iPSC-derived neurons (iNeurons) abrogated uptake of aSyn fibrils. Moreover, exogenous expression of *GPNMB* enabled HEK293 cells to internalize aSyn fibrils, a property this cell line does not possess at baseline^4^. In the context of malignancy and inflammation, the GPNMB ECD can be shed from the cell surface to mediate non-cell-autonomous effects^15,17^. Concordant with these reports, we also observed that transient transfection of *GPNMB* into HEK293 cells conferred ability to internalize fibrillar aSyn to all cells in the culture, regardless of whether an individual cell expressed *GPNMB* highly or not. These observations led us to hypothesize that GPNMB can act non-cell-autonomously to enable aSyn fibril uptake **(Figure 1A)**.

**Figure 1.**
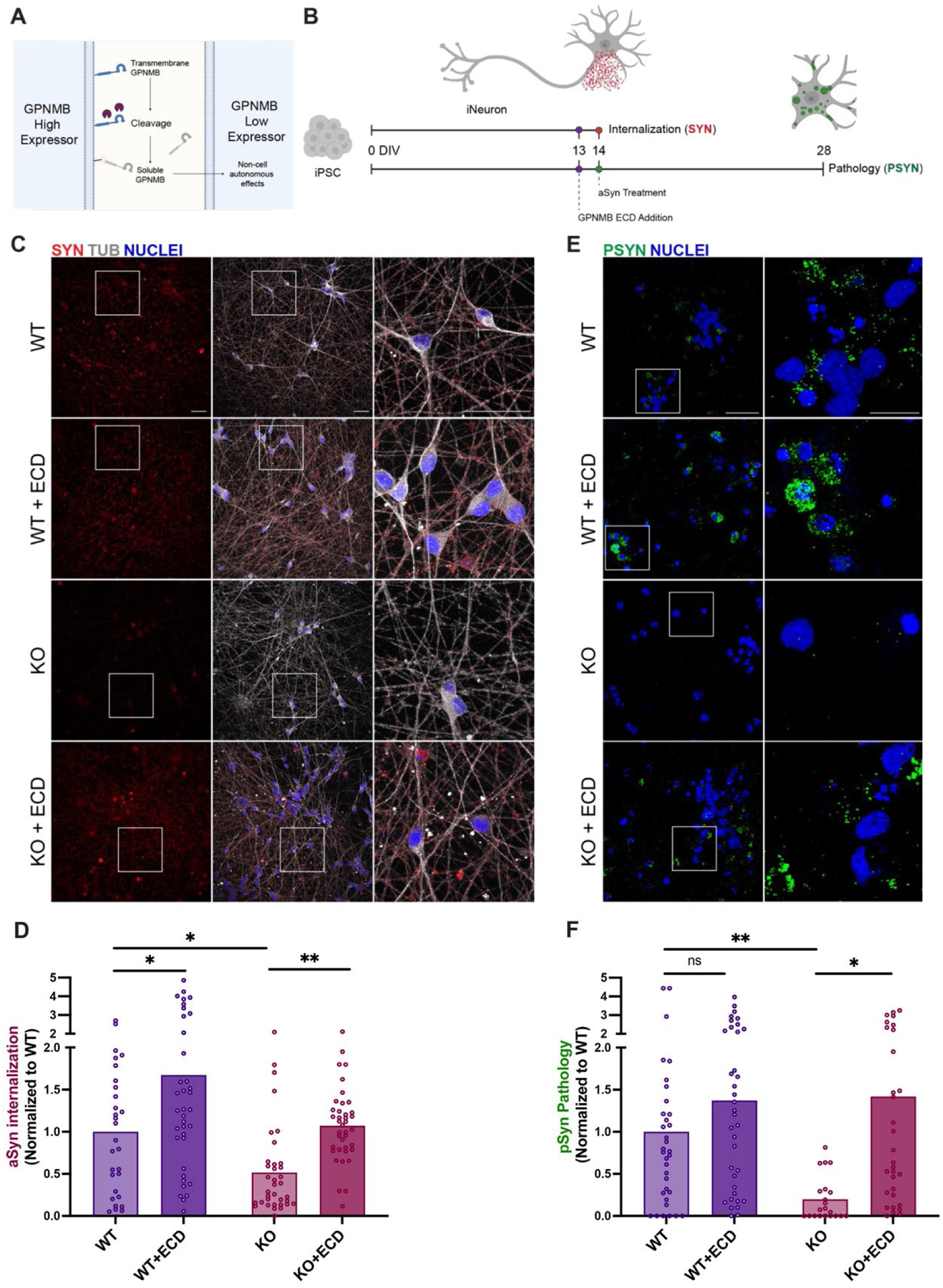
The soluble GPNMB extracellular domain (ECD) enhances fibrillar aSyn uptake and aggregation in iNeurons. **(A).** Proposed mechanism of GPNMB ECD action. The GPNMB ECD is cleaved from the membrane by metalloproteases, where it can act non-cell-autonomously to trigger downstream molecular events, including enhanced fibrillar aSyn uptake. **(B).** Experimental scheme encompassing differentiation of iNeurons, addition of GPNMB ECD and aSyn fibrils, and timing of aSyn fibril uptake and phosphorylated aSyn pathology assessments. **(C).** Representative images of WT iNeurons treated with labeled aSyn PFFs alone (WT) or PFFs and GPNMB ECD (WT + ECD); or KO iNeurons treated with labeled PFFs alone (KO) or PFFs and GPNMB ECD (KO + ECD). Nuclei were stained with DRAQ5. Representative image (scale bar = 50um) with inset highlighting region of interest (scale bar = 20um). PFFs internalized by cells are shown in red, while tubulin staining (Tub) is shown in grey. **(D).** Quantification of aSyn puncta in internalization experiments. KO neurons exhibit a significant reduction in internalization compared to WT neurons (\**p*=0.04). This is rescued by treating KO neurons with GPNMB ECD (\*\**p*=0.007). Treatment of WT neurons with GPNMB ECD also results in significantly increased internalization of labeled aSyn fibrils (\**p*=0.017). Bar depicts mean. Each dot represents one field, with 4-10 fields quantified per replicate (well, n = 6) across 3 differentiations. Statistics were calculated using nested t-test (one-tailed, given expected direction) to account for non-independent fields (see Methods). **(E).** Representative images of WT iNeurons treated with untagged aSyn PFFs alone (WT) or PFFs in combination with GPNMB ECD (WT + ECD); and KO iNeurons treated with untagged PFFs alone (KO) or in combination with GPNMB ECD (KO + ECD). aSyn aggregates phosphorylated at serine 129 (pSyn, green) were detected 14 days after PFF treatment, following extraction of soluble proteins with 1% Triton-X, using an antibody against phosphorylated synuclein (pS129, EP1536Y). Nuclei were stained with DRAQ5. Representative image (scale bar = 50um) with inset highlighting region of interest (scale bar = 20um). **(F).** Quantification of insoluble, phospho-Syn (pSyn) aggregates, normalized to WT. GPNMB KO neurons exhibit a significant reduction in aggregate number compared to WT neurons (\*\**p*=0.007) and to KO neurons treated with GPNMB ECD (\**p*=0.038). In contrast, GPNMB ECD treatment did not significantly alter pSyn aggregates in WT neurons compared to baseline. Bar depicts mean. Each dot represents one field, with 3-7 fields quantified per replicate (well, n = 6) across 4 differentiations. Statistics were calculated using nested t-test (one-tailed, given expected direction) to account for non-independent fields (see Methods).

To determine whether the GPNMB ECD can act non-cell-autonomously on neurons, we differentiated induced pluripotent stem cells (iPSC) into cortical neurons (iNeurons) (**Figure 1B**), deriving iNeurons with wild-type (WT) levels of *GPNMB* expression as well as isogenic *GPNMB* knockout (KO) iNeurons, created by genome editing of a parental iPSC-line as previously described^28^. In agreement with our prior report, loss of *GPNMB* in iNeurons reduced cellular uptake of aSyn fibrils when compared to WT iNeurons (p=0.04; **Figure 1C**, **1D**). However, addition of human recombinant GPNMB ECD to the cell culture medium of KO iNeurons rescued their ability to internalize aSyn (p=0.007; **Figure 1C**, **1D**). Treatment of WT neurons with GPNMB ECD also resulted in significantly increased internalization of labeled aSyn fibrils (p=0.017; **Figure 1C**, **1D**).

We confirmed the effects of exogenous addition of the GPNMB ECD in a PD model in which aSyn fibrils are used to seed alpha-synuclein (aSyn) pathology in iNeurons (**Figure 1B**). As previously reported, WT iNeurons form insoluble, hyperphosphorylated aSyn aggregates 14 days after one-time seeding with aSyn fibrils, while *GPNMB* KO iNeurons form minimal aSyn aggregates (p=0.007; **Figure 1E**, **1F**). Remarkably, however, treatment of *GPNMB* KO iNeurons with the GPNMB ECD added to the culture medium alone was sufficient to convert them to a WT neuron phenotype (p=0.038; **Figure 1E**, **1F**).

Finally, we tested for direct interaction between the GPNMB ECD and aSyn fibrils in an *in vitro* context. In pulldown assays, using the GPNMB ECD as bait robustly captured aSyn fibrils, but not aSyn monomer, confirming that the ECD fragment of GPNMB is sufficient for interaction and that the GPNMB ECD preferentially binds fibrillar forms of aSyn (**Figure S1**).

Taken together, these data demonstrate that the GPNMB ECD can act in a non-cell-autonomous manner to enhance internalization of aSyn fibrils in human neurons, impacting subsequent formation of aSyn pathology.

### GPNMB is widely expressed in the human brain with highest expression in microglia

Previous studies have reported GPNMB protein expression in several areas of the central nervous system (CNS), including in the spinal cord in ALS^20^, frontal white matter in Nasu-Hakola disease^19^, hippocampus and frontal cortex in neuro-HIV^22^, and substantia nigra in PD^23^. Additionally, single-cell transcriptomic studies of human brain^23^ and animal models^29^ support *GPNMB* expression in multiple CNS cell types. However, a comprehensive analysis of GPNMB expression by brain region and cell type within the human brain is lacking.

To address this gap, we characterized postmortem cases from the Penn Integrated Neurodegenerative Disease Brain Bank (**Table S1**), performing immunohistochemical staining of GPNMB across multiple brain regions (cingulate, temporal cortex, hippocampus, cerebellum) from neurologically normal controls (NCs, n = 3-6 for each brain region) and individuals with a neuropathological diagnosis of Lewy body disease (LBD, the neuropathological correlate of clinical PD and the related disorder Dementia with Lewy bodies (DLB), n = 13-15 for each brain region). In both NC and LBD cases, we found widespread GPNMB expression across all brain regions assessed (**Figure 2A**), with a non-significant trend towards increased expression in LBD individuals (**Figure 2B**). Moreover, a subset of LBD cases studied had concomitant AD pathology. However, the presence or absence of concomitant AD pathology did not significantly affect GPNMB expression levels (**Figure 2B**).

**Figure 2.**
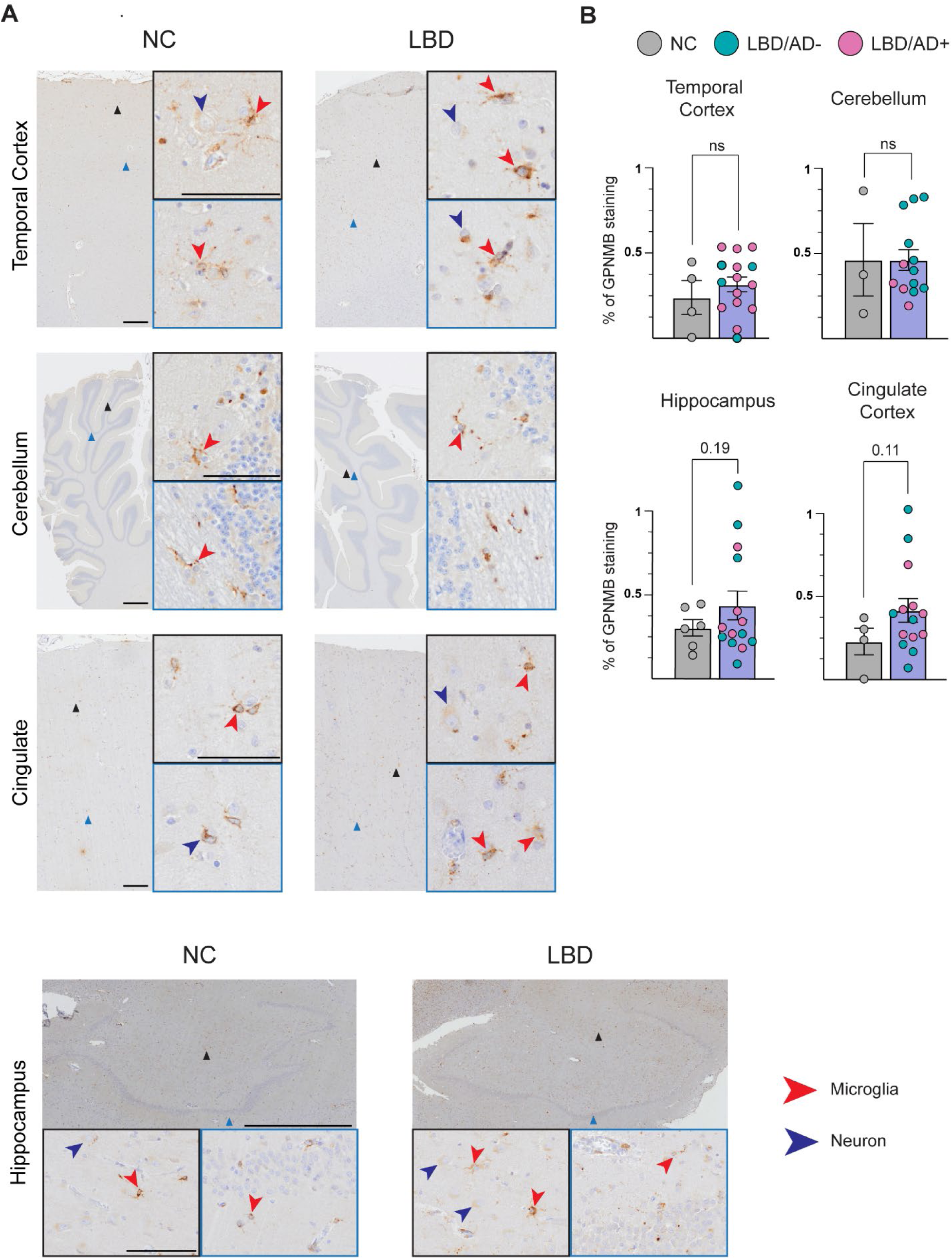
Expression of GPNMB in the human brain, across brain regions and neuropathological diagnoses. **(A).** Representative immunohistochemistry images of GPNMB stained in the temporal cortex, cerebellum, cingulate cortex, and hippocampus across neurologically normal control (NC), and Lewy body disease (LBD) cases. GPNMB is detected (AF2550 antibody) in cells with the morphology of neurons (blue arrowheads) and microglia (red arrowheads) across all brain regions. Representative image (scale bar = 2mm) with insets highlighting region of interest (scale bar = 100um), indicated on low-power image with triangle corresponding to color of inset frame. **(B).** Quantification of GPNMB protein expression (percentage area with GPNMB staining) in the temporal cortex, cerebellum, hippocampus, and cingulate cortex across NC, LBD cases. Within the LBD group, the presence (LBD/AD+, pink dots) or absence (LBD/AD-, teal dots) of concomitant AD pathology is indicated. Bars depict mean +/- SEM, with each dot representing one case. Statistics were calculated using an unpaired t-test (two-tailed).

In our neuropathological cases, we observed GPNMB expression in cells with neuronal morphology and in cells with microglial morphology. To further understand cell-type specific *GPNMB* expression patterns, we integrated and analyzed data from four previously-reported single cell RNA-sequencing (scRNAseq) studies of PD and NC brain^23,30–32^ (**Table S2**). For the three midbrain studies, microglia were the cell type most likely to express *GPNMB* (**Figure 3A, 3B, 3C**). Moreover, microglia expressing *GPNMB* were increased in PD compared to NC (p=0.0067, **Figure 3D**). Indeed, analyzing microglia from all four studies together, or from the three scRNAseq studies of midbrain together, we found that *GPNMB* is one of the top genes differentiating PD vs. NC microglia (**Figure 3E, Figure S2**, **Table S3** and **Table S4**). In contrast, analyses of neurons or astrocytes from the same scRNAseq studies showed no significant differences in proportions expressing *GPNMB* comparing PD to NC brain (**Figure S3**).

**Figure 3.**
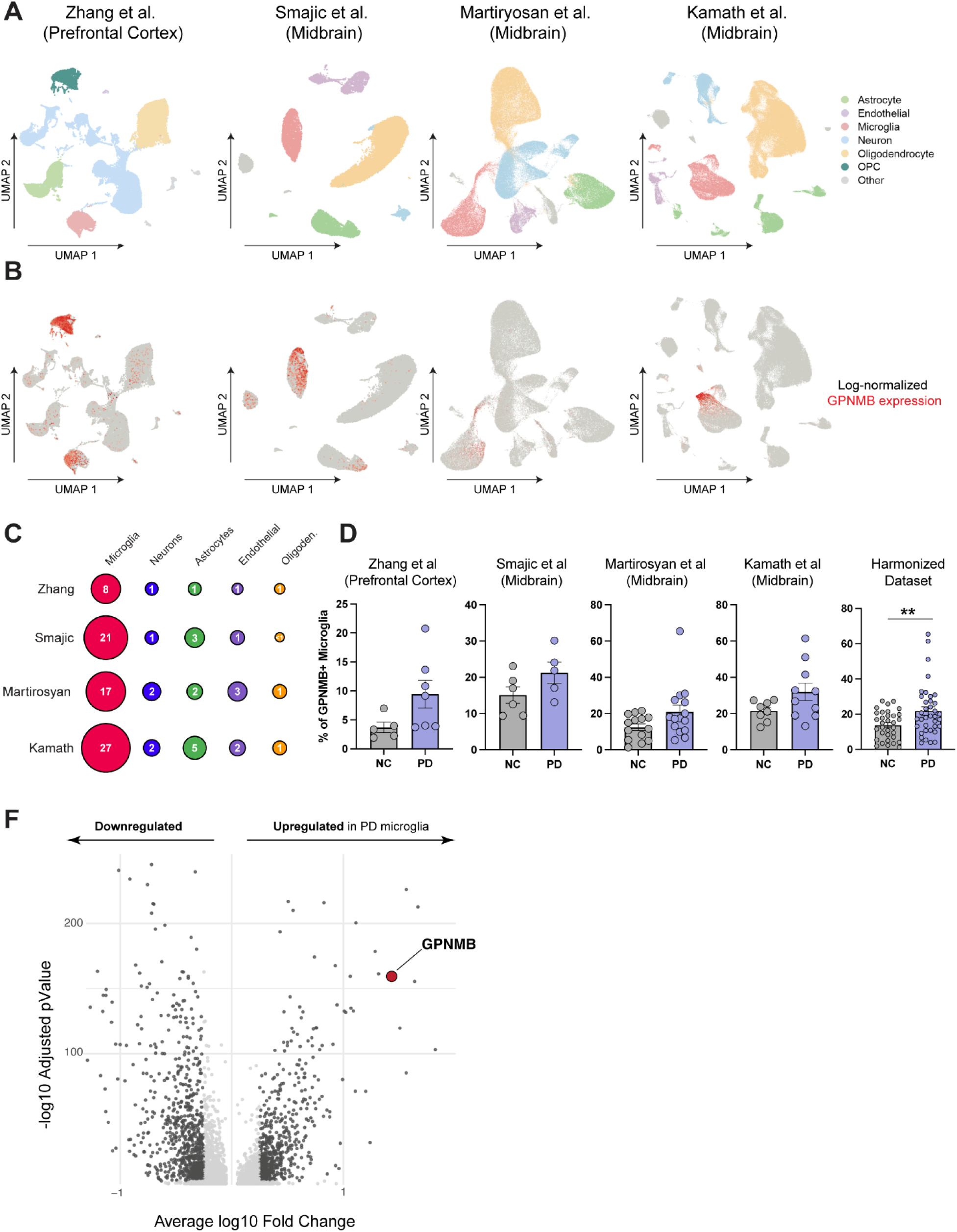
*GPNMB* is predominantly expressed in microglia, and PD brain has more *GPNMB*-expressing microglia. **(A).** UMAP projections of four single-cell RNA sequencing (scRNAseq) datasets. Cell types are assigned to clusters according to canonical markers (see Methods). **(B).** Corresponding UMAP projections showing expression of *GPNMB* transcript, visualized as normalized log-transformed counts (red). *GPNMB* expression is enriched in the microglia cell cluster across all datasets examined. **(C).** Percentage of cells expressing detectable levels of *GPNMB* transcript, subdivided by cell type (neurons, astrocytes, oligodendrocytes, endothelial cells, microglia – see Methods for assignment), from each scRNAseq study. **(D).** Percentage of microglia expressing detectable levels of GPNMB transcript, stratified by neuropathological diagnosis (NC or PD) from each scRNAseq study, as well as the integrated data from all four studies, showing increased numbers of GPNMB+ microglia in PD (\*\**p*=0.0067). Statistics were calculated using an unpaired t-test (two-tailed). **(E).** Volcano plot comparing microglia from PD vs. NC samples in the integrated dataset. *GPNMB* is one of the most upregulated genes in PD microglia.

We confirmed these scRNAseq data at the protein level with double-label immunofluorescence staining of human brain sections. We found GPNMB protein expression in cells that expressed the neuronal marker NeuN, as well as cells that expressed the microglial marker Iba1 (**Figure 4A**, **4B**).

**Figure 4.**
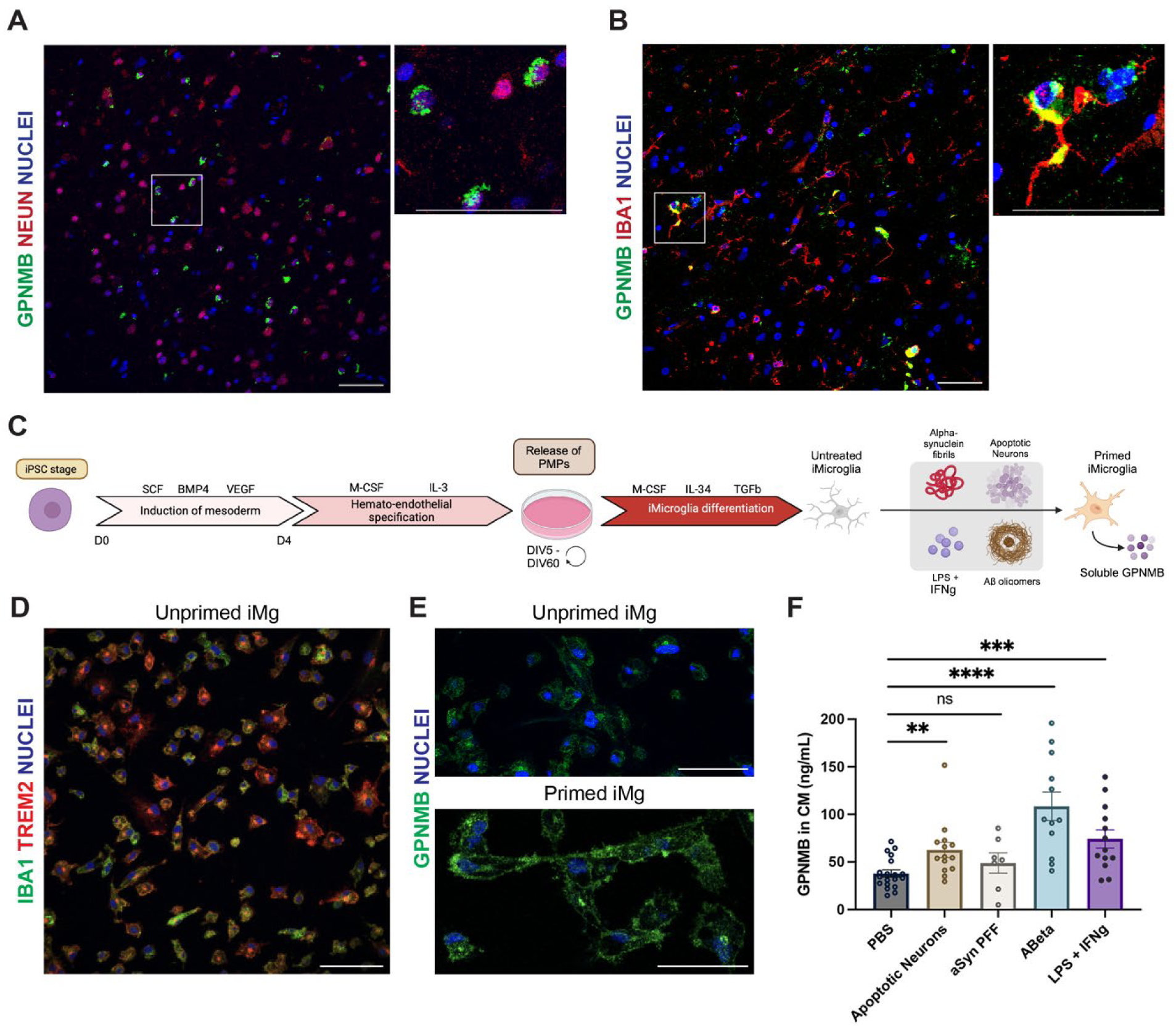
iMicroglia release GPNMB into medium. **(A and B).** Double-label immunofluorescence staining of human brain tissue demonstrates GPNMB (green) in cells expressing neuronal markers (NeuN, red, panel A) and in cells expressing microglial markers (Iba1, red, panel B). Scale bar = 50uM. **(C).** Differentiation of iMicroglia from iPSCs and schematic of treatments. **(D).** Differentiated iMicroglia stained with microglial markers Iba1 (Abcam-5076, green) and TREM2 (D8I4C, red). (**E and F**). Intensity of GPNMB staining (sc-271415, green) increases when iMicroglia are primed with liposaccharide/interferon-gamma. Scale bar = 50uM. **(F).** GPNMB protein measured by ELISA in the conditioned medium of iMicroglia cultures increases when iMicroglia are treated with apoptotic neurons (135k/cm^2^), beta-amyloid oligomers (1uM), and lipopolysaccharide (100ng/mL) and interferon gamma (20ng/mL), but not when iMicroglia are treated with aSyn PFFs (10ug). Statistics were calculated using unpaired two-tailed t-tests. **p<0.01, ***p<0.001, ****p<0.0001.

These findings demonstrate that *GPNMB* is widely expressed in human brain tissue in neurons as well as microglia. However, *GPNMB* expression is highest in microglia, and particularly in microglia from PD brain.

### Microglia-derived GPNMB increases in response to neurodegeneration and enhances uptake of aSyn fibrils in human neurons

An emerging literature supports upregulation of *GPNMB* by microglia and macrophages in response to various stimuli, including neurodegeneration-related insults^25^. Having observed high *GPNMB* expression in microglia, and particularly in microglia from PD brain (**Figure 3**, **Figure 4B**), we asked whether microglia might secrete GPNMB, and whether GPNMB secretion would increase in response to neurodegeneration-related triggers.

In lysates from the HMC3 microglial cell line, we detected a dose-dependent increase in GPNMB protein when cells were exposed to apoptotic neurons (ANs, **Figure S4**). Moreover, in conditioned medium (CM) from iPSC-derived microglia differentiated according to previously published protocols^33^ (iMicroglia, **Figure 4C**, **4D**, **4E, Figure S5**), we detected GPNMB protein at baseline, which increased when iMicroglia were exposed to ANs (p=0.005), amyloid-beta oligomers (p<0.0001), and lipopolysaccharide/interferon-gamma (p<0.001), but not to aSyn fibrils (**Figure 4F**).

Having demonstrated that iMicroglia release GPNMB into the surrounding medium at baseline and in response to various stimuli, we tested CM from iMicroglia for ability to induce uptake of aSyn fibrils in WT and *GPNMB* KO iNeurons. Because GPNMB secretion by iMicroglia in response to stimuli is variable in degree, we used CM from untreated iMG. Application of CM from WT iMicroglia (CM^WT^) fully rescued uptake of aSyn fibrils in *GPNMB* KO iNeurons (p=0.021; **Figure S6A** and **S6B**) and increased baseline aSyn uptake in WT iNeurons (p=0.049; **Figure S6A** and **S6B**). Additionally, application of CM^WT^ enabled formation of insoluble, hyper-phosphorylated aSyn aggregates in *GPNMB* KO iNeurons (p=0.003; **Figure 5A** and **5B**) and increased formation of aSyn aggregates in WT iNeurons (p=0.042). Importantly, application of CM from *GPNMB* KO iMicroglia (CM^KO^) did not have this effect (**Figure 5A** and **5B**), and direct testing of GPNMB levels from CM^KO^ showed no detectable GPNMB (**Figure S4C** and **S4D**), confirming that GPNMB is the decisive factor promoting development of aSyn aggregates.

**Figure 5.**
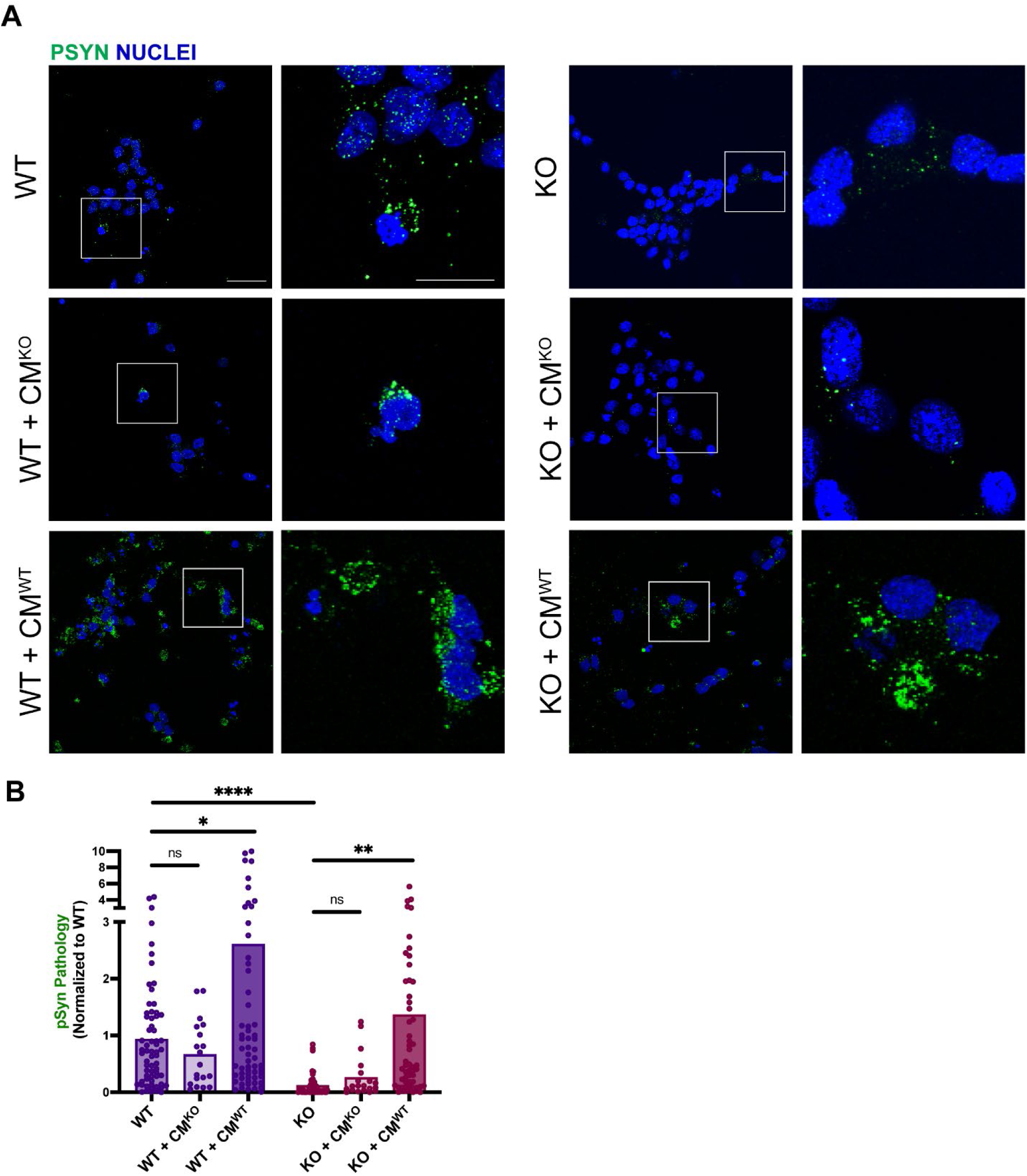
Conditioned media from iMicroglia enhances aSyn uptake in iNeurons. **(A).** Left: Representative images of iPSC-derived neurons (iNeurons) including wild-type non-treated (WT), WT treated with CM collected from KO iMicroglia (WT + CM^WT^), and WT treated with CM collected from KO iMicroglia (WT + CM^KO^). Right: Representative images of iNeurons including GPNMB knock-out non-treated (KO), KO treated with CM collected from KO iMicroglia (KO + CM^KO^), and KO treated with CM collected from WT iMicroglia (KO + CM^WT^). 14 days after seeding with untagged aSyn PFFs, serine-129-phosphorylated aSyn aggregates resistant to extraction with 1% Triton-X are stained with EP1536Y antibody (green). Nuclei stained with DRAQ5 in blue. Scale bars: 50 μm (overview), 20 μm (inset). **(B)** Quantification of aSyn aggregates in neurons exposed to CM from both WT and KO iMicroglia. KO iNeurons show decreased insoluble pSyn aggregates when compared to WT iNeurons (\*\*\*\**p* <0.0001). Both WT iNeurons (*\*p=*0.042) and KO iNeurons (*\*\*p=*0.003) treated with CM^WT^ increased formation of insoluble pSyn aggregates, but CM^KO^ does not have this effect. Each dot represents one field, with 3-7 fields per replicate (well, n=8) across 4 differentiations, and bar depicts the mean. Statistics were calculated using nested t-test (one-tailed, given expected direction of effect) to account for non-independent fields (see Methods).

Taken together, our results show that iMicroglia release GPNMB in the presence of neurodegeneration-related stimuli, and that GPNMB released from iMicroglia is sufficient to enable aSyn uptake and pathology formation in *GPNMB* KO iNeurons.

### GPNMB-mediated fibrillar aSyn uptake can be blocked by anti-GPNMB monoclonal antibodies

In the preceding sections, we have demonstrated that microglia release GPNMB in response to ANs, and that GPNMB can act non-cell-autonomously to enhance neuronal uptake of aSyn fibrils, resulting in development of more aSyn pathology. We next turned our attention to therapeutic development, asking whether this pathophysiological cascade can be blocked with antibodies targeting the GPNMB ECD.

To do this, we raised 42 monoclonal antibodies (mAbs) against the GPNMB ECD and tested them for ability to block internalization of aSyn fibrils. We first screened our antibodies for ability to block aSyn fibril uptake in HEK293 cells engineered to stably express GPNMB-GFP (**Figure 6A**). This HEK293-GPNMB system allows for rapid assessment of labelled aSyn fibril uptake (**Figure 6B**), in the presence of low and high doses of each mAb (**Figure 6C**). 15 of the 42 anti-GPNMB mAbs blocked aSyn fibril uptake in this initial screen, with 4 blocking uptake at both low and high doses (**Figure 6D, Figure S7, S8**).

**Figure 6.**
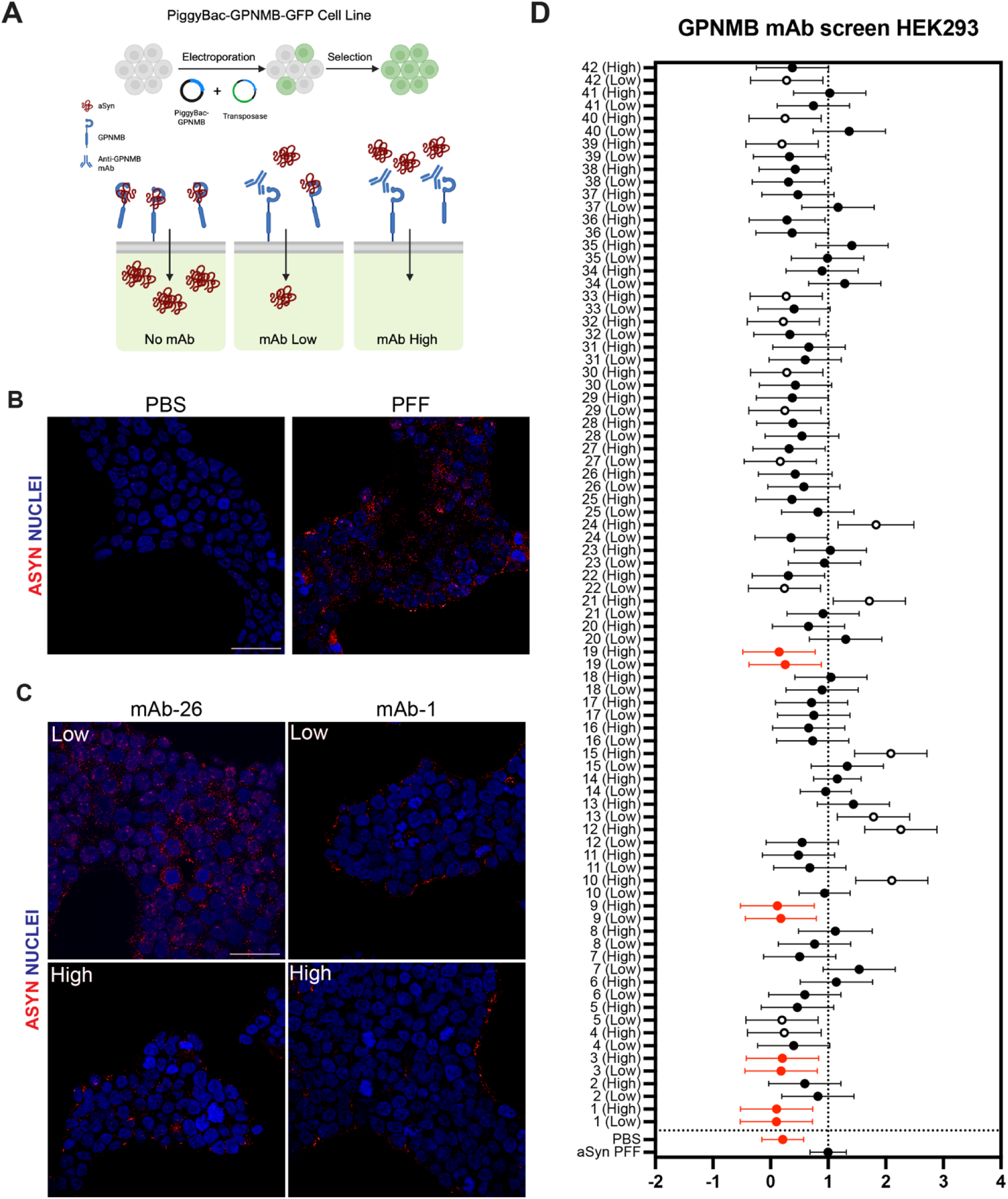
Cellular uptake of aSyn fibrils is blocked by treatment with antibodies targeting the GPNMB ECD. **(A).** HEK293 cells were engineered to stably express GPNMB-GFP, and cells were treated with labelled aSyn PFFs in the presence or absence of monoclonal antibodies targeting the GPNMB ECD. **(B).** Representative immunofluorescence images showing aSyn uptake in GPNMB-GFP-expressing cells under control (PBS) or aSyn fibril (PFF) treatment. Constitutive expression of GPNMB-GFP is sufficient to drive uptake of labeled aSyn fibrils (red). Nuclei labeled with DRAQ5 (blue). Scale bar = 50um. **(C).** Representative images from cells treated with two anti-GPNMB mAbs of varying effect. In the case of mAb-26 (left), treatment with antibody, particularly at high doses, trended towards blocking uptake of labeled aSyn fibrils (red), while mAb-1 (right) significantly blocked uptake of aSyn fibrils at high (750 ng/mL) or low (75 ng/mL) concentrations. Nuclei labeled with DRAQ5 (blue). Scale bar = 50um. **(D).** 42 different clones of mAbs raised against the GPNMB ECD were tested for ability to block uptake of aSyn PFFs in HEK293-GPNMB cells. Four anti-GPNMB mAbs (mAb-1, mAb-3, mAb-9, mAb-19, highlighted in red) significantly decreased aSyn PFF uptake at both doses tested (*FDR-corrected *p*<0.05). Additional anti-GPNMB mAb clones (clear circles) significantly altered aSyn PFF uptake for one dose, but high and low doses did not show consistent effects. For each antibody dose, 5 fields of view were collected from two replicate wells. Means and 95% confidence intervals for antibody effects are shown, normalized to the no-antibody, aSyn PFF-seeded condition. Statistics were calculated using a nested one-way ANOVA comparing each antibody dose with PFF-only condition, with FDR (Benjamini-Krieger-Yukitieli) set at 0.05.

We next asked whether anti-GPNMB mAb treatment could also block aSyn fibril uptake and subsequent development of aSyn pathology in iNeurons. We tested two mAbs (mAb-1 and mAb-26) with varying levels of blocking effect in the initial screen. We found that both mAb-26 and mAb-1 blocked development of aSyn pathology in iNeurons at high doses, with mAb-1 blocking development of aSyn pathology at low doses as well, recapitulating findings from the HEK293 screen (**Figure 7**, **Table S5**). In contrast, isotype-matched non-targeting IgG antibodies showed no blocking effect (**Figure 7**). To verify that mAb-1 blocks development of aSyn pathology by binding to the GPNMB ECD, we incorporated additional control conditions, where GPNMB ECD or CM from WT iMicroglia (CM^WT^) was added to the culture medium of *GPNMB* KO iNeurons, and mAb-1 was tested for effect. In this context, mAb-1 again blocked subsequent development of aSyn pathology (p<0.05 for both GPNMB ECD and CM^WT^ conditions, **Figure S9**). Furthermore, treating *GPNMB* WT iNeurons with mAb-1 reduced formation of aSyn pathology even in the presence of CM from WT iMicroglia (p=0.018, **Figure S9**).

**Figure 7.**
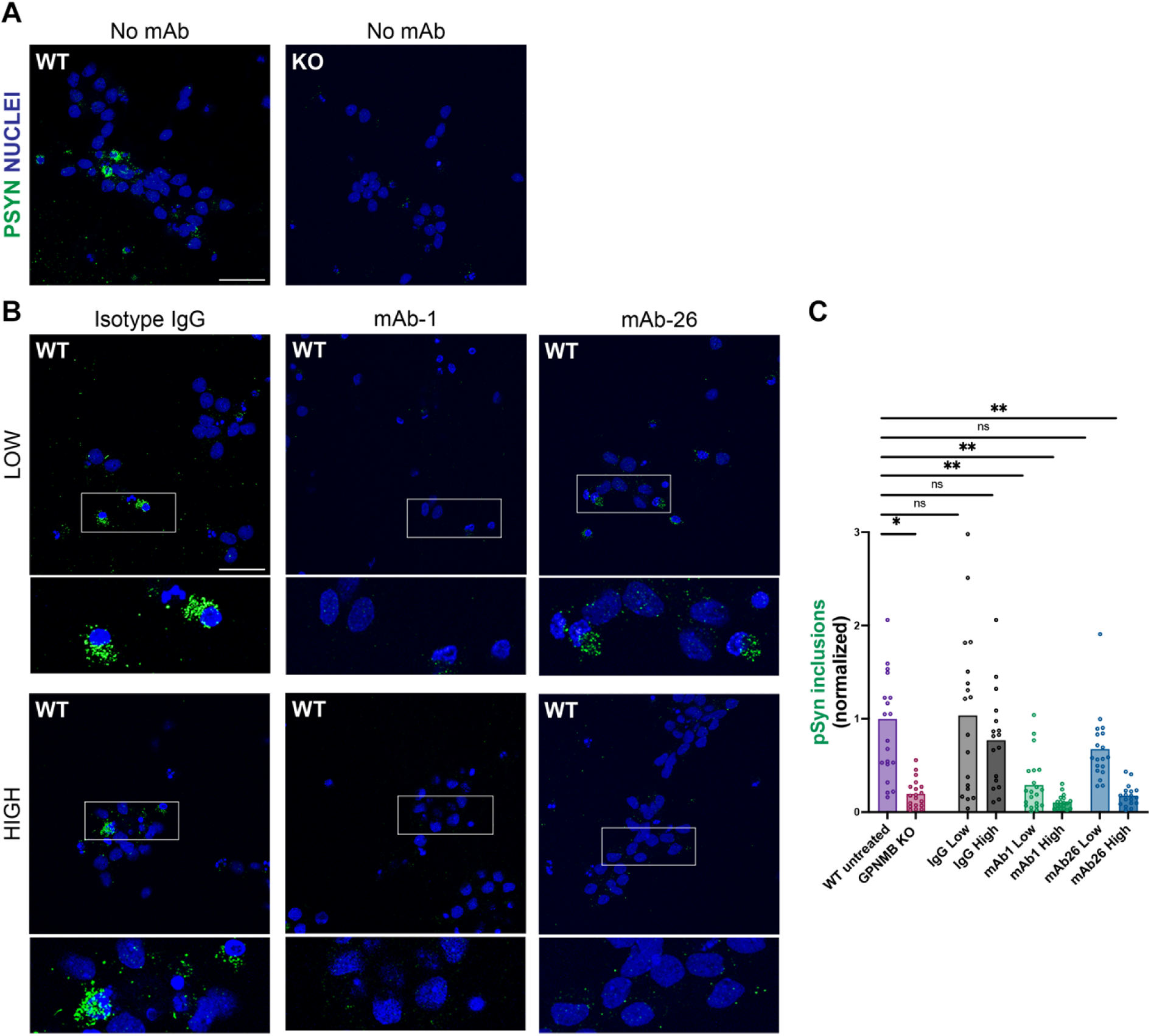
Anti-GPNMB mAb treatment rescues iNeurons from developing aSyn pathology. **(A).** Control conditions showing formation of hyperphosphorylated insoluble aSyn aggregates in WT iNeurons, but not GPNMB KO iNeurons, 14 days after seeding with aSyn PFFs. Phospho-serine 129 aSyn aggregates (green) were stained with EP1536Y, and nuclei with DRAQ5. Scale bar = 50 μm. (**B**) Representative images of WT iNeurons seeded with aSyn fibrils and also treated with low (top row, 75 ng/mL) or high (bottom row, 750 ng/mL) dose of negative control (isotype-matched IgG) antibody, anti-GPNMB mAb-26, or anti-GPNMB mAb-1. Cells were stained for hyperphosphorylated, 1% Triton-X-insoluble aSyn aggregates (green, EP1536Y) and nuclei (DRAQ5, blue) 14 days after seeding with aSyn PFFs. Scale bar = 50 μm. **(B).** Quantification of phosphorylated αSyn aggregates in WT iNeurons seeded with PFF and treated with negative control antibody (IgG, grey), anti-GPNMB mAb-1 (green), or anti-GPNMB mAb-26 (blue). Untreated WT iNeurons (purple) and GPNMB KO iNeurons (pink) seeded with PFF are also shown for reference. Each dot represents one field, with 3-6 fields per replicate (n=4 replicate wells), across 2 differentiations. Statistics were calculated using nested one-way ANOVA, followed by pairwise comparisons of each antibody and dose with PFF-only condition, adjusted for testing 3 antibodies (see Methods). One-tailed p-values are presented, given expected direction of effect. Treatment with mAb-1 at low (adjusted *p*=0.004), and high (adjusted *p*=0.001) doses, as well as mAb-26 at high dose (adjusted *p*=0.003) significantly rescued pSyn pathology.

These results demonstrate that a subset of anti-GPNMB mAbs can block aSyn fibril uptake by neurons, rescuing them from subsequent development of aSyn pathology.

### Genotypes associated with *GPNMB* expression predict spread of aSyn pathology in human brain tissue

Our experiments suggest that increased *GPNMB* expression, genetically encoded or triggered by microglial exposure to neurodegeneration-related insults, leads to increased neuronal uptake of aSyn fibrils. Increased cellular uptake of aSyn fibrils, in turn, could lead to more aggressive cell-to-cell spread of aSyn pathology throughout the brain. To test this expectation, we genotyped 1675 neurodegenerative disease cases at rs199347, a single nucleotide polymorphism (SNP) we had previously demonstrated to associate with levels of *GPNMB* expression^4^ (Cases described in **Table S6**). We asked whether rs199347 genotypes predicted spread of aSyn pathology, ranging from no aSyn pathology (score = 0), to amygdala-only aSyn pathology (score = 1), to brainstem-predominant aSyn pathology (score = 2), to transitional/limbic aSyn pathology (score = 3), to diffuse/neocortical aSyn pathology (score = 4), as captured by the McKeith Criteria for Lewy Pathology stage^34–36^.

As shown in **Figure 8A**, *GPNMB* genotypes at rs199347 associated significantly with the extent of Lewy pathology (p=0.010). Individuals homozygous for the PD risk allele (AA, associated with higher *GPNMB* expression) had the most widespread aSyn pathology (Mckeith stage 1.31 ± 0.07), followed by heterozygous individuals (GA: McKeith stage 1.21 ± 0.06), and those homozygous for the protective allele (GG: McKeith stage 1.00 ± 0.08, (**Figure 8B**). In contrast, rs199347 genotypes did not associate with extent of beta-amyloid or tau pathology (**Figure 8C** and **8D**).

**Figure 8.**
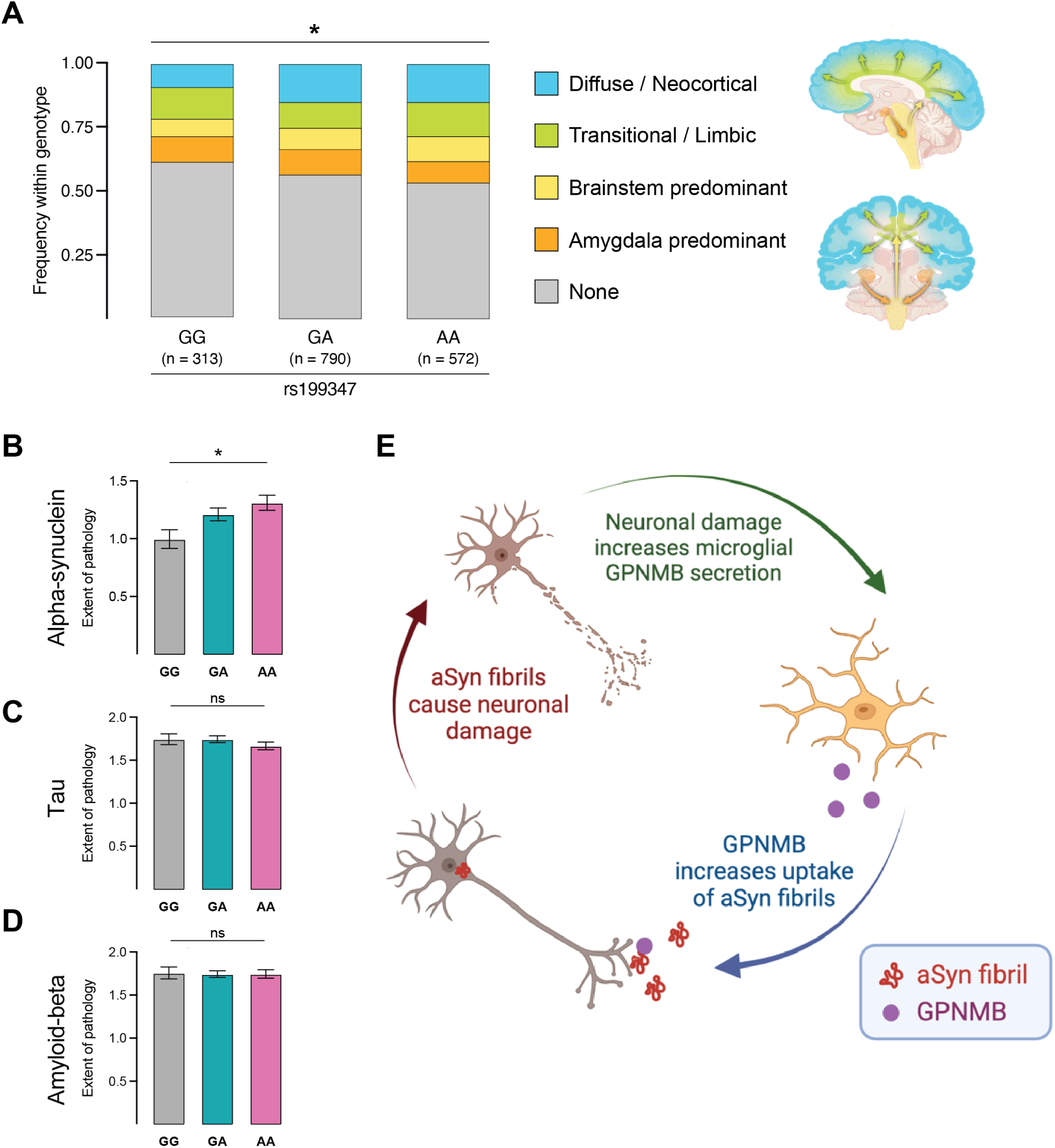
Association of rs199347 genotypes with extent of aSyn pathology. **(A).** Stacked bar plots representing the distribution of Lewy body pathology stages across rs199347 genotypes (GG, GA, AA, where the A allele associates with PD risk and higher *GPNMB* expression) in 1675 postmortem cases analyzed neuropathologically (n=344 LBD, n=626 AD, n=661 non-LBD/AD neurodegenerative disease, n = 44 normal aging, based on primary neuropathological diagnosis, see **Table S6** for details). Lewy body pathology is classified as diffuse/neocortical, transitional/limbic, brainstem-predominant, amygdala-predominant, or none. Rs199347 genotype effect on extent of Lewy body pathology assessed in a linear regression adjusted for age at death and sex (\**p*=0.010). **(B–D)**. Extent of alpha-synuclein (**B**), tau (**C**), and amyloid-beta (**D**) pathology across *GPNMB* genotypes for n=1675 cases. Extent of pathology is captured by McKeith stage for alpha-synuclein, Braak stage for tau, and CERAD stage for amyloid-beta. Mean stage ± SEM is shown. For each pathology, rs199347 genotype effects were assessed using a nonparametric test (Kruskal-Wallis), followed by pairwise comparisons between each genotype. Rs199347 genotype groups differed significantly for extent of alpha-synuclein pathology (\**p*=0.018), with pairwise differences for GG vs. AA carriers (*p*=0.015), but not for tau or amyloid-beta pathology. **(E).** Proposed model of GPNMB function in microglia-neuron interactions in PD pathogenesis. Following exposure to apoptotic neurons or other neurodegeneration-related insults, microglia upregulate *GPNMB* expression and increase secretion of GPNMB. GPNMB can then act on surrounding cells, such as neurons, to facilitate uptake of pathological forms of aSyn, contributing to the spread of aSyn pathology. Development of aSyn pathology further damages neurons, triggering microglia to release more GPNMB. Figure schematics created in Biorender.

## DISCUSSION

Following our 2022 report that *GPNMB* is the target gene of a common variant PD risk locus identified by genome-wide association^4^, and that GPNMB is critical to the cellular uptake of aSyn fibrils, we present evidence here for a non-cell-autonomous mechanism by which GPNMB enhances development of PD pathology, alongside a potential method for therapeutic blockade. Specifically, using cellular models of PD pathogenesis, we demonstrate that the non-membrane-anchored GPNMB ECD – whether added alone as recombinant protein or secreted from microglia – enhances cellular uptake of pathological forms of aSyn and subsequent development of aSyn pathology in neurons. Using human neuropathological specimens, scRNAseq data, and iMicroglia cell culture, we further demonstrate that GPNMB is expressed in both neurons and microglia across multiple brain regions, with higher expression in microglia, and that microglia secrete GPNMB in response to neurodegeneration. These data suggest a model in which GPNMB participates in a positive feedback loop between microglia and neurons, where microglia secrete GPNMB in response to neuronal damage, enhancing uptake of pathological forms of aSyn in neurons, causing more neuronal damage, triggering more GPNMB secretion by microglia (**Figure 8E**). An intriguing possibility is that microglial GPNMB secretion, triggered by cortical insults, may even drive spread of neuronal aSyn pathology from the brainstem to limbic and cortical areas of the brain, with implications for the development of clinical symptoms such as dementia. Importantly, we also demonstrate that antibodies targeting GPNMB may be able to interrupt this pathophysiological cascade. Finally, by analyzing 1675 neuropathological cases, we show that carriers of genotypes associated with higher *GPNMB* expression have more widespread Lewy pathology, underlining the relevance of this target to human disease.

Several aspects of our study warrant emphasis. First, evidence for the non-cell-autonomous effects of GPNMB ECD comes from multiple forms of GPNMB (recombinant and microglia-derived), across multiple cellular models (microglial cell lines and iMicroglia), increasing confidence. Second, support for our overall framework comes from both manipulative experiments in neuronal and microglial models, allowing us to dissect effects, as well as extensive human neuropathological data, giving depth to what would otherwise be reductionist models. In this regard, we highlight the fact that we previously linked rs199347 genotype to GPNMB protein levels in human CSF and human plasma, in samples from >700 individuals with and without *GBA1* mutations, demonstrating that PD risk genotypes consistently associate with higher GPNMB expression, important for interpreting the mechanism of common variant associations with PD at this locus^37^. Third, our monoclonal antibody data provides evidence that the observed effects on cellular uptake of aSyn fibrils are specific to GPNMB and, importantly, offers an immediate avenue for therapeutic development^38^.

Limitations of our study should be considered alongside its strengths. For example, while the cellular models presented here allow for precise manipulation of each cell type, with the ability to gauge ensuing molecular phenotypes, *in vivo* manipulation of *GPNMB* in all cell types, or in neurons or microglia selectively, would add to our understanding of disease pathophysiology. Moreover, our demonstration that effects on aSyn fibril uptake can be achieved by a non-membrane-tethered GPNMB ECD raises the question of what cellular receptors may mediate GPNMB’s effects. Receptors previously reported for GPNMB include CD44^18^, syndecan-4^39^, and subunits α1 and α3 of the Na+/K+ ATPase^40–42^, while FAM171A2 and LRP1 have been reported as neuronal receptors involved in the internalization of aSyn fibrils^43,44^. Studies investigating these candidates, or non-receptor-mediated avenues of cellular uptake, could add to our understanding of GPNMB’s role in PD pathogenesis. Third, the finding that microglia secrete GPNMB in response to various neurodegeneration-related triggers raises the question of what functions, aside from increasing aSyn fibril uptake in neurons, secreted GPNMB may have in modulating neuroinflammatory cascades. Indeed, a recent study reports that microglial cell lines increase their uptake of multiple disease proteins in response to GPNMB^45^. Finally, while we focused our work on microglia and neurons, investigating GPNMB’s actions in additional cell types (*e.g.* oligodendrocyte precursor cells and astrocytes) could add to our understanding.

While these open questions offer valuable future directions, we note that total loss of GPNMB in humans results in only a mild dermatological phenotype – autosomal-recessive amyloidosis cutis dyschromica^46^. This suggests that strategies to reduce GPNMB or block its function may be well-tolerated. With more than 10 million people worldwide affected by PD, our data support GPNMB as an attractive target for disease-modifying therapy.

## Data Availability

All data produced in the present study are available upon reasonable request to the authors.
All analysis scripts can be found on the listed github page.

https://github.com/Chen-PlotkinLab/Harmonized_PD_Dataset.

## ACKNOWLEDGEMENTS AND FUNDING

We thank the many patients and their families for contributing biosamples to this research. This study was supported by the NIH (R37 NS115139, P30 AG010124, U19 AG062418, and PO1 AG084497) as well as SPARK-NS. Work in the Chen-Plotkin laboratory is additionally supported by the Parker Family Chair and the Lipman Family Fund.

## KEY RESOURCES TABLE

| **ANTIBODIES** |  |  |
| --- | --- | --- |
| REAGENT or RESOURCE | DILUTION /<br>CONCENTRATION | IDENTIFIER |
| Anti-alpha-synuclein antibody (MJFR1), IF 1:300 | (IF) 1:300 | Ab138501 (Abcam) |
| Anti-alpha-synuclein S129 antibody (EP1536Y) | (IF) 1:1000 | Ab51253 (Abcam) |
| Anti-beta-III tubulin antibody | (IF) 1:1000 | ab52623 (Abcam) |
| Anti-TREM2 antibody (D8I4C) | (IF) 1:1000 | 91068 (Cell Signaling) |
| Anti-Iba1 antibody | (IF) 1:1000 | Ab5073 (Abcam) |
| Anti-GAPDH antibody | (WB) 1:3000 | 2-RGM2 (Advanced Immunochemical) |
| Anti-GPNMB antibody | (IF) 1:250 | sc-271415 (Santa Cruz) |
| Anti-GPNMB antibody | (WB) 1:1000; (IHC) 1:250 | AF-2550 (R&D) |
| Goat Anti-Rabbit Alexa Fluor 488 | (IF) 1:1000 | A-21206 (Invitrogen) |
| Goat Anti-Mouse Alexa Fluor<br>594 | (IF) 1:1000 | A-21203 (Invitrogen) |
| Donkey Anti-Goat Alexa<br>Fluor 647 | (IF) 1:1000 | A-21447 (Invitrogen) |
| DRAQ5 | (IF) 1:1000 | 62551 (ThermoFisher) |
| Goat Anti-Mouse HRP | (WB) 1:3000 | Jackson Immuno Research<br>(115-035-062) |
| Biotinylated Rabbit Anti-Goat | (IHC) 1:5000 | BA-5000-1.5 (Vector<br>Laboratories) |
| Biotinylated Anti-GPNMB<br>sc-271415 Antibody | (NP) 0.001 ug/mL | sc-271415 (Santa Cruz),<br>biotinylated in-house |
| Biotinylated Anti-Alpha-Synuclein syn211 Antibody | (NP) 0.001 ug/mL | ab80627 (Abcam),<br>biotinylated in-house |
| Biotinylated Mouse IgG1, $\kappa$<br>Isotype Control | (NP) 0.001 ug/mL | 349040 (BD Bioscience) |
| Peroxidase Streptavidin | (NP) 1:1000 | 016-030-084 (Jackson) |

IF: Immunofluorescence ; WB: Western blot; IHC : Immunohistochemistry, NP: Nickel Plate

CELL LINES
| NAME | SPECIES | OBTAINED FROM |
| --- | --- | --- |
| iNGN | Homo sapiens | George Church lab, Harvard Medical School<br>(PMID: 25403753) |

| OTHER REAGENTS |  |  |
| --- | --- | --- |
| NAME | COMPANY | PRODUCT ID |
| Recombinant human<br>GPNMB ECD | Abcam | Ab151652 |
| CHAPS hydrate | Sigma | C3023 |
| Paraformaldehyde, EM Grade,<br>Purified | Electron Microscopy<br>Sciences | 30525-89-4 |
| Poly-D-lysine | Gibco | A3890401 |
| PBS | Gibco | 14190-136 |
| Pierce 660nm Protein assay<br>reagent | Thermo Scientific | 22660 |
| 4-20% MiniProtean TGX gels | MiniProtean | FB2489 |
| 0.2um nitrocellulose<br>membrane | BioRad | 162-0112 |
| Neurobasal A medium | ThermoFisher | 10888022 |
| DMEM-F12 medium | Gibco | 11320033 |
| DMEM-F12 KO medium | Gibco | 10829018 |
| mTESR Plus Basal Medium | StemCell Technologies | 100-0276 |
| ReleSR passaging solution | StemCell Technologies | 100-0483 |
| Accutase dissociation reagent | StemCell Technologies | 07920 |
| B27 Supplement (50X) | ThermoFisher | 17504044 |
| N2 Supplement (100X) | ThermoFisher | 17502048 |
| MEM non-essential amino<br>acid | ThermoFisher | 11140050 |
| Glutamax (100X) | Gibco | 35050061 |
| Penicillin-Streptomycin | Gibco | 15070063 |
| Recombinant human BDNF | StemCell Technologies | 78005.1 |
| Recombinant human NT3 | StemCell Technologies | 78074.1 |
| Recombinant murine laminin | R&D Systems | 3837-A4-050 |
| Doxycycline hyclate | Sigma | D9891-5G |
| Y-27632 | Tocris | 1254 |
| Essential 8 Medium | Gibco | A1517001 |
| X-VIVO 15 Serum-free<br>Hematopoietic Cell Medium | Lonza Bioscience | 02-053Q |
| 2-Mercaptoethanol | Gibco | 21985023 |
| Recombinant human BMP4 | Peprtech | 120-05ET-100UG |
| Recombinant human SCF | Peprtech | 300-07-100UG |
| Recombinant human VEGF | Peprtech | 100-20-100UG |
| Recombinant human IL-3 | Peprtech | 200-03-100UG |
| Recombinant human M-CSF | Peprtech | 300-25-100UG |
| Recombinant human IL-34 | Peprtech | 200-34-100UG |
| Recombinant human TGFb | Peprtech | 100-21-10UG |
| GEM21 Neuroplex<br>Supplement | GeminiBio | 400-160-010 |
| AF594 and untagged human aSyn PFFs | CNDR, University of Pennsylvania |  |
| Beta-Amyloid oligomers (1-42) | Anaspec | AS-20276 |
| Lipopolysaccharides (LPS) | Cell Signaling Technology | 14011 |
| Human Interferon-γ (hIFN-γ) Recombinant Protein | Cell Signaling Technology | 80385 |
| Pierce Nickel Coated ELISA Plates | ThermoFisher | 15442 |
| Human aSyn monomer | CNDR, University of Pennsylvania |  |
| Human GPNMB Protein, His Tag | Acro Biosystems | GPB-H5229-100ug |
| di-Sodium hydrogen phosphate dihydrate (Na<sub>2</sub>HPO<sub>4</sub>•2H<sub>2</sub>O) | MilliporeSigma | 1.06580.0500 |
| Sodium phosphate monobasic (NaH<sub>2</sub>PO<sub>4</sub>) | Sigma-Aldrich | S3139-500G |
| Sodium Chloride (NaCl) | Fisher Chemical | S671-500 |
| Imidazole | Thermo Scientific | A10221.36 |
| Tween-20 | BioRad | 1706531 |
| Bovine Serum Albumin | Sigma-Aldrich | A3059-100G |
| EZ-Link™ Sulfo-NHS-LC-Biotin, No-Weigh™ Format | Thermo Scientific | A39257 |
| Water | Sigma-Aldrich | 270733-1L |
| Slide-A-Lyzer™ MINI<br>Dialysis Devices, 10K<br>MWCO | Thermo Scientific | 69574 |
| 1-Step™ TMB ELISA<br>Substrate Solutions | Thermo Scientific | 34028 |
| Phosphoric Acid | LabChem | LC186501 |

## METHODS DETAILS

### Maintenance of iPSC Cultures

Human induced pluripotent stem cells (iPSCs) expressing doxycycline-inducible neurogenin-1 and neurogenin-2 (iNGN1, iNGN2) were cultured on Cultrex-coated 6cm tissue culture dishes in mTeSR™ Plus medium supplemented with 5×mTeSR™ Plus supplement (StemCell Technologies). Cultures were maintained with medium changes every 48 hours and passaged when ∼80% confluent using StemMACS™ XF passaging solution (StemCell Technologies) or ReLeSR™ (StemCell Technologies). iPSCs were kept at all times at 5% O2 hypoxic conditions.

### Neuronal Differentiation

When ∼80% confluent, iPSCs were dissociated into a single-cell mixture using Accutase™ (StemCell Technologies) and centrifuged at 300xg for 5 minutes. Cells were then plated at a density of 700,000 cells/well onto Cultrex-coated 6-well plates in N2 pre-differentiation medium containing KnockOut™ DMEM/F12 (Gibco) supplemented with 1×N-2 supplement (EMSCO/Fisher), 1×MEM Non-Essential Amino Acid solution (EMSCO/Fisher), 10 ng/mL brain-derived neurotrophic factor (BDNF; EMSCO/Fisher), 10 ng/mL neurotrophin-3 (NT-3; EMSCO/Fisher), 0.2 μg/mL laminin (EMSCO/Fisher), 2 μg/mL doxycycline hyclate (Sigma), and 10 μg/mL Y-27632 (Tocris). Medium was refreshed 24 hours after plating with Y-27632 to increased cell adhesion and again at 48 hours without Y-27632. Pre-differentiated iPSCs were kept at all times at 5% O2 hypoxic conditions.

Round 12mm glass coverslips were coated with 10 μg/mL poly-D-lysine (PDL; Sigma) prepared in borate buffer (pH 8.4) and incubated for 1 hour at room temperature in the dark. The PDL-coated coverslips were then washed three times with tissue-culture-grade water (Lonza) and coated with Cultrex for 1 hour at room temperature or stored at 4C until use.

Pre-differentiated iPSCs at 72hr after induction were dissociated with Accutase, centrifuged at 300xg for 5minutes, and resuspended in neuronal differentiation medium containing 50% DMEM/F12, 50% Neurobasal-A (Invitrogen), 1× MEM Non-Essential Amino Acid solution, 0.5× N-2 supplement, 0.5× GlutaMAX™ supplement (Invitrogen), 0.5× B27™ supplement (Invitrogen), 10 ng/mL BDNF, 10 ng/mL NT-3, 1 μg/mL laminin, and 2 μg/mL doxycycline. Cells were seeded at a density of 200,000 cells/well onto PDL/Cultrex-coated coverslips in 12-well plates for immunofluorescence analysis. Half-medium changes were done at specific intervals to maintain optimal culture conditions. Seven days after induction (Day 7, D7), half of the medium was removed and replaced with fresh neuronal differentiation medium without doxycycline. On D14, an additional half-volume of fresh medium was added without removing the existing medium. For experiments requiring 28 days of incubation, another half-volume of fresh medium was added on D21. iNeurons were kept at all times at normoxia conditions.

### Microglial Differentiation

iPSCs were differentiated to iMicroglia following an adaptation from an existing protocol^33^. iPSCs were cultured in a 6-well format pre-coated with Matrigel (Corning). On day 0 (D0), iPSCs were dissociated into small clumps using TrypLE Express and resuspended in ice-cold Essential 8 (E8) Media (ThermoFisher). Cells were counted and plated at 10,000 cells per well into 96-well ultra-low attachment plates (Corning #7007) in 150uL EB media containing E8 medium supplemented with BMP4 (50 ng/mL), SCF (20 ng/mL) and VEGF (50 ng/mL) as well as a Rock inhibitor. Plates were centrifuged at 1,000 rpm for 5 minutes and incubated in a hypoxia chamber. On D2, 150uL of EB media was added to each well. On D4, EBs (32-40 per well) were gently captured using wide-bore tips and transferred to 10cm dishes pre-coated with Matrigel and containing 12 mL of hematopoietic myeloid medium (HMM). HMM medium was prepared using X-VIVO 15 (Lonza), 55 μM 2-mercaptoethanol, M-CSF (100 ng/mL), and IL-3 (25 ng/mL). The dishes were placed in a low oxygen incubator and gently shaken to evenly distribute the EBs. Media changes were performed at regular intervals: 12 mL of HMM was added on day 8, media was replaced on day 12, and an additional 12 mL was added on day 16. By day 20, floating HMPs were harvested. PMPs were collected every 8 days and plated in uncoated dishes at a density of 750 cells/mL in HMM for expansion. PMP populations were characterized using flow cytometry for CD45 and CD11b markers, ensuring >70% double-positive cells. Progenitors were plated at 125,000 cells per well in 12-well plates in microglia maturation medium (MMM). MMM was prepared using Neurobasal-A medium (ThermoFisher) supplemented with Gem21 without vitamin A (1X), GlutaMAX (1X), IL-34 (100 ng/mL), M-CSF (100 ng/mL), and TGFβ (50 ng/mL). Media was changed on D4 and D8. Mature iMicroglia expressing microglia-specific markers were observed by D7, and cells were used for downstream experiments on D10. All reagents and their sources are detailed in the Key Resources Table.

### α-Synuclein Pre-Formed Fibril (PFF) Internalization Experiments

Human wild-type pre-formed fibrils (PFFs) and Alexa Fluor 594 (AF594)-labeled α-synuclein (aSyn) PFFs were provided by Dr. Kelvin Luk. AF594-labeled PFFs were generated by spiking unlabeled αSyn assembly reactions with AF594-labeled αSyn (1:9 w/w) before fibril initiation^47^. PFFs were stored at −80°C until use. Stocks (5 mg/mL) were diluted in DPBS to a working concentration of 0.1 mg/mL and sonicated using a Diagenode Bioruptor® Plus (30 seconds on/off, high power, 10 minutes total, 10°C). Sonicated PFFs were added to neuronal differentiation media at a final concentration of 10 μg/mL (1 mL media/well in a 12-well plate) and applied to cells. For conditions subjected to recombinant GPNMB ECD (rGPNMB) treatment, iNeurons were pre-treated with 100ng/mL of rGPNMB (Abcam) for 24hr before PFF addition. For experiments involving monoclonal anti-GPNMB antibody treatment (generated as described below), iNeurons were pre-treated 24 hours prior to PFF addition with either a low or high dose of the antibody. The low dose corresponded to a 1:10 molar ratio of IgG to PFFs (750ng per condition), while the high dose corresponded to a 1:1 molar ratio (7.5ug per condition), assuming ∼200KDa as the molecular weight of PFFs. Cells were subsequently processed as above. Each PFF internalization experiment was repeated independently three times from iNeurons corresponding to independent differentiations, with two technical replicates (wells) per condition. For internalization assays, iPSC-derived neurons (iNeurons) were incubated with PFFs at 4°C for 30 minutes, followed by an additional 1.5 hours at 37°C, while kept at all times in the dark. Cells were then quenched with Trypan blue^48^ to remove fluorescence signal from extracellular PFFs, fixed and stained.

To induce aSyn pathology in aggregation experiments (PSYN), neuronal differentiation media was replaced at D14 with medium containing unlabeled human PFFs at a concentration of 1ug/mL (1mL/well in a 12-well plate). iNeurons were incubated with PFFs for 14 days with a half-medium addition at D21, as described above. Pre-treatment with GPNMB ECD or anti-GPNMB mAb were performed as above. For measurement of hyperphosphorylated insoluble aSyn inclusions, protein soluble extraction was performed by fixing coverslips in 2% paraformaldehyde and 1% Triton X-100 in dPBS. For blocking, 3% bovine serum albumin in dPBS without saponin was used. For experiments using recombinant GPNMB ECD (rGPNMB), iNeurons were pre-incubated with 100 ng/mL rGPNMB (Abcam) for 24 hours prior to the addition of PFFs. Each PFF aggregation experiment was repeated independently three times from iNeurons corresponding to independent differentiations, with two technical replicates (wells) per condition.

### Generation of a GPNMB-PiggyBac Construct and Stable Line

The GPNMB-PiggyBac construct (Pb-CAG-GPNMB-GFP-WPRE-SV40) was generated by cloning the RG207615 construct (Origene) into a modified PiggyBac Vector (Addgene #110824, provided by Dr. Stewart Anderson’s lab, UPenn). Cloning was achieved via PCR amplification to introduce 5’ NheI and 3’ PmeI restriction sites using the following primers:

- F: TAAGCAGCTAGCGCCACCATGGAATGTCTCTACTATTTCCTG
- R: CGGCCGTTTAAACTCTTTCTTCAC

The PiggyBac transposase construct (pCAG-PBase) was obtained from the Anderson lab (UPenn) and corresponds to Addgene #54285. HEK293 cells were dissociated using trypsin, counted, and plated at 700,000 cells per well in a 6-well format and allowed to grow overnight. The following morning, cells were transfected using 4 micrograms of GPNMB PiggyBac and PBase constructs and 10 microliters of lipofectamine in 1mL of serum-free DMEM. This was placed in an incubator at 37C with 5% CO2 for 5 hours.

After 5 hours, the transfection media was removed and cells were given pre-warmed complete culture medium for recovery. After recovery, cells were plated sparsely onto 15cm^2^ plates and monitored daily. Once colonies formed, individual colonies were picked and expanded in culture medium supplemented with blasticidin (50 μg/mL) to select for stable GPNMB expression. Clonal populations were maintained and expanded for downstream experiments.

### Anti-GPNMB monoclonal antibody generation and screening

Monoclonal anti-GPNMB antibodies (mAbs) targeting the extracellular domain of human GPNMB were generated by immunization of 5 BALB/c mice three times each with recombinant human GPNMB ECD (AA 22-486). Post-immunization bleeds were tested for antibody response to GPNMB via western blotting and immunofluorescence using transiently-transfected PiggyBac-GPNMB cells. For generating hybridomas, spleens from four mice were fused to create two fusion pools. The 42 most promising hybridoma clones were selected for downstream characterization (mAb-1 to mAb-42). All 42 clones were confirmed to recognize GPNMB by ELISA (**Table S5**).

For immunofluorescence, PiggyBac-GPNMB cells were plated at densities of 100,000 cells per well, on poly-D-lysine (PDL)-coated 12 mm glass coverslips in a 12-well plate format. Similar to above, cells were pre-treated 24 hours prior to PFF addition with either a low or high dose of each of the mAbs. The low dose corresponded to a 1:10 molar ratio of IgG to PFFs (75ng per condition), while the high dose corresponded to a 1:1 molar ratio (750ng per condition), assuming ∼200KDa as the molecular weight of PFFs. Cells were subsequently processed as above.

### Imaging processing and quantification

Monochrome images were acquired in Z-stacks measuring 0.25um per slice using a Leica SP5 confocal microscope equipped with a 40×oil immersion objective. Details of antibody concentrations are provided in the Key Resources Table.

Quantification of PFF internalization and phosphoSyn pathology was performed using a semi-automatic image processing pipeline. Z-stack images comprising 15 slices (0.25 μm per slice) were processed to select the central 10 slides, and a maximum intensity projection was generated. These projection files were subsequently analyzed using CellProfiler with a custom-built pipeline that measured 1) mean aSyn fluorescence per cell; 2) number of aSyn- or phosphoSyn-positive puncta (aSyn Dots); and 3) total nuclei count. For each field captured, the number of puncta was normalized to the number of nuclei.

### Immunoblotting

Samples were collected from cells using RIPA buffer and prepared for immunoblotting by diluting them in 4x Laemmli Sample Buffer (Bio-Rad Laboratories). Samples were directly loaded into 4-20% polyacrylamide TGX gels (Bio-Rad Laboratories). Proteins were separated by electrophoresis and transferred onto 0.2 μm nitrocellulose membranes (Bio-Rad Laboratories). Membranes were blocked in 5% non-fat milk prepared in TBS-T for 1 hour. Membranes were incubated overnight at 4C with primary antibodies, as detailed in the key resources table. After washing, membranes were incubated with HRP-conjugated secondary antibodies for 2 hours at room temperature, extensively washed using TBS-T and TBS, and developed using Clarity Max Western ECL substrate (Bio-Rad Laboratories).

### RNA extraction, Reverse Transcription, and Quantitative PCR

RNA was extracted from cells using RNeasy Plus Kit (Qiagen), and RNA purity was measured with a NanoDrop spectrophotometer. Purified RNA (1 μg) was treated with DNase I (ThermoFisher) for 15 minutes at room temperature, followed by inactivation with 2 μL of 25 mM EDTA and heating at 65C for 10 minutes. Following DNase treatment, RNA was mixed with random hexamers and a 10 mM dNTP mix. The mixture was incubated at 65C for 5 minutes and chilled on ice for at least 1 minute. Reverse transcription was performed by adding 5X buffer, water, 0.1 M DTT, and Super Script Reverse Transcriptase (ThermoFisher) to the RNA/primer mixture. The reaction was incubated at 25C for 5 minutes, followed by 50 minutes at 50C, and terminated at 70C for 15 minutes. The resulting cDNA was diluted with 120 μL of nuclease-free water. Quantitative PCR (qPCR) was performed using the ΔΔCt method and the following primers:

- GPNMB (F): 5’- CTTCTGCTTACATGAGGGAGC-3’
- GPNMB (R): 5’- CTCCCTTCCAGGAGTTTTTCC-3’
- ACTB: Applied Biosystems, Cat# 4352935E

### Enzyme-linked immunosorbent assay (ELISA)

GPNMB levels in conditioned media (CM) and brain lysates were quantified using the Human Osteoactivin/GPNMB DuoSet ELISA kit (R&D Systems, #DY2550) in conjunction with the DuoSet ELISA Ancillary Reagent Kit (R&D Systems, #DY008B). CM from PiggyBac-GPNMB HEK293 cells was diluted 1:500, while CM from iMicroglia was diluted 1:25. ELISA was performed according to the manufacturer’s instructions, absorbance was measured at 450 nm using a microplate reader, with correction at 570 nm to account for background noise. Standard curves were generated using the provided GPNMB standards to calculate absolute concentrations of GPNMB in the samples. Measurements were performed in technical duplicates to ensure reproducibility.

### Immunohistochemistry

Formalin-fixed, paraffin-embedded tissue samples from the hippocampus, temporal cortex, cerebellum and cingulate cortex were obtained from the brain bank of the Penn Center for Neurodegenerative Research (CNDR). Sections (6 μm thick) were deparaffinized in xylenes and rehydrated through a descending ethanol series (100%, 95%, 80%, and 70%). Endogenous peroxidase activity was quenched by incubating sections in a solution of 70% methanol and 30% H₂O₂ for 30 minutes. Antigen retrieval was performed by heating the slides in citric acid-based Antigen Unmasking Solution (Vector Laboratories) at 95°C for 30 minutes in a water bath. Following cooling, sections were rinsed in TBS (0.1 M Tris Buffer) and blocked for 5 minutes with blocking buffer containing TBS-T, 2% fetal bovine serum (FBS), and 3% bovine serum albumin (BSA). Sections were incubated overnight at 4°C with a primary anti-GPNMB antibody as described in the Key Resources Table. After washing in TBS, sections were incubated at room temperature for 1 hour with a biotinylated secondary antibody (Vector Laboratories, 1:1000). Signal amplification was performed using the VECTASTAIN ABC Standard Kit (Vector Laboratories) for 1 hour, and staining was visualized with ImmPACT DAB (Vector Laboratories) for 30 seconds. Sections were counterstained with Harris Hematoxylin (Thermo Scientific) for 50 seconds, followed by dehydration through an ascending ethanol series (70%, 80%, 95%, 100%) and xylenes. Coverslips were mounted using Cytoseal (Thermo Scientific), and allowed to dry overnight. Slides were scanned using an Olympus VS200 slide scanner. Stained tissue was analyzed using HALO software (Indica Labs) to calculate the percentage of strong staining using a manually-defined threshold in the annotated ROI. The percentage of positively stained area was automatically calculated.

### scRNA-seq Data Processing and Analysis

Single-cell RNA sequencing raw data were downloaded from the datasets listed in **Table S2** and processed using the Seurat package in R. Broadly, data containing barcodes, gene features, and expression matrices were extracted and verified for completeness and converted into Seurat objects. Quality control was performed to remove low-quality cells and artifacts, filtering out cells with fewer than 200 genes, more than 3,000 genes, or greater than 15% mitochondrial gene content. Data normalization was conducted using the Seurat package^49^, specifically the LogNormalize() method, followed by scaling and identification of 2000 highly variable genes. Dimensionality reduction was performed using Principal Component Analysis (PCA), and significant principal components were selected based on the elbow plot method, ranging from 15 for individual datasets to 50 PCs for the merged datasets after integration. Uniform Manifold Approximation and Projection (UMAP) was applied for visualization of the data in two-dimensional space (**Supplementary Figure 2**). Cell clustering was performed using a shared nearest neighbor (SNN) modularity optimization-based algorithm. Cell types were annotated manually by contrasting selected cell type markers with the following canonical genes:

- Microglia cluster: *CSF1R, P2RY12, TMEM119, CX3CR1, CD74*
- Neuron cluster: *SYT1, SNAP25, GRIN1*
- Astrocyte cluster: *AQP4, GFAP, SLC1A3*
- Endothelial cluster: *CLDN5, VWF, FLT1, PECAM1*
- Oligodendrocyte cluster: *MBP, MOG, PLP1, MOBP*

Results were reported as percentage of cells expressing GPNMB and data was stratified by patient diagnosis according to the clinical data provided by each study.

To generate an integrated atlas of human microglia across studies, the Harmony package^50^ was used to correct for batch effects across datasets. Two final objects were created: one including only midbrain datasets (**Figure S2**) and another combining midbrain and cortical datasets (**Figure 3**). For each integrated object, differential expression testing between PD and NC groups was performed using Seurat’s FindMarkers() function, which uses the non-parametric Wilcoxon’s Sum Rank test. Volcano plots were generated using the resulting tables of differentially expressed genes, filtered by a log fold change threshold of 0.05 and a minimum expression fraction of 5% (**Tables S3 and S4**). All analysis scripts can be found on https://github.com/Chen-PlotkinLab/Harmonized_PD_Dataset.

### Association of rs199347 genotype with neuropathological staging

Linear regression analysis (Pathology extent ∼ rs199347 + sex + age at death) was performed to assess the association between rs199347 genotype and the extent of alpha-synuclein pathology as classified by the McKeith criteria, adjusting for sex and age at death (**Figure 8A**). In assessing multiple different pathologies (**Figure 8B**), Kruskal-Wallis non-parametric tests were performed to assess the relationship between rs199347 genotype and alpha-synuclein pathology (McKeith), amyloid-beta (CERAD and Thal), and tau pathology (Braak). Neuropathological and demographic information was collected from a dataset of 1675 postmortem neuropathological cases from the Center for Neurodegeneration Research (CNDR) at the University of Pennsylvania. rs199347 genotypes were determined via SNPtype assays designed using the D3 Assay Design Tool (Standard BioTools, San Francisco, CA). The brain bank lacks olfactory bulb data for most individuals, so assessment of olfactory bulb pathology was excluded from the McKeith Staging. To harmonize between 6-stage and 3-stage Braak tau scoring (employed by the Penn CNDR brain bank at different historical periods), cases staged with Braak 0-6 scores were converted to Braak 0 to 3 scores as follows: Braak 0 was converted to 0, Braak I - II were converted to Braak 1, Braak III-IV were converted to Braak 2, and Braak V – VI were converted to Braak 3. Covariates for the analysis included sex and age at death. Patient demographics can be found in **Table S6**.

### Preparation of amyloid-beta

Amyloid-beta (1-42) oligomers were prepared by dissolving the lyophilized peptide (Anaspec) in 1% NH4OH, followed by dilution to the working stock concentration in 1X PBS. The solution was then incubated at 4C for 24 hours to allow for oligomer formation.

### Quantification and Statistical Analysis

Statistical testing was performed as indicated in the figure legends. Cell biological data were quantified for aSyn PFF internalization or phosphoSyn pathology using CellProfiler (see *Imaging processing and quantification*). For experiments in which each biological replicate (well or coverslip) had multiple technical replicates (fields captured), nested ANOVA or t-test was used to account for interdependent data (fields within a well) in order to test for significance of genotype (GPNMB WT or KO) or treatment effects. For antibody effect in HEK293 screen (**Figure 6**), all antibody treatments were compared to aSyn PFF-only condition, with adjustment for multiple hypothesis testing at an FDR of 0.05 using the two-stage linear step-up procedure of Benjamini, Krieger, and Yekuteli. For antibody effect in iNeurons (**Figure 7**), all antibody treatments were compared to aSyn PFF-only condition, with adjustment for testing 3 antibodies (nominal p-value x 3). Graphs were generated and analyses performed in Prism.

No statistical methods were used to predetermine sample sizes. In experiments involving treatments with monoclonal antibodies, investigators were blinded during data collection for at least one biological replicate. Further statistical details can be found in the figure legends.

### Antibody Biotinylation Protocol

Antibodies for the nickel plate protein-protein interaction assay were biotinylated using EZ-Link™ Sulfo-NHS-LC-Biotin, No-Weigh™ Format (Thermo Scientific) according to the manufacturer’s guidelines. Briefly, a 1 mg vial of biotin was equilibrated to room temperature, then diluted with 180 uL of HPLC-grade water (Sigma-Aldrich) to a working concentration of 10 mM biotin. The 10 mM biotin was incubated for two hours at 4°C with end-over-end agitation. 35-fold molar excess biotin was added to each antibody and the biotin-antibody mixture was incubated for three hours at 4°C with end-over-end agitation. Unbound biotin was removed using Slide-A-Lyzer™ MINI Dialysis Devices (Thermo Scientific) according to the manufacturer’s guidelines. The tube was filled with 1.2 mL of PBS (Gibco) and the biotin-antibody mixture was added to the dialysis cup. The device was incubated for two hours at 4°C with orbital shaking. Following the incubation, the tube was rinsed once with 1.5 mL of PBS and was filled with 1.2 mL of PBS. The device was incubated again for two hours at 4°C with orbital shaking, after which the dialyzed antibody was collected from the cup, aliquoted, and stored at -20°C prior to use. Biotinylation was performed for the following antibodies: anti-GPNMB (sc-271415), anti-alpha-synuclein (syn211), and Mouse IgG1, κ Isotype Control antibody (BD Biosciences).

### Nickel Plate Protein-Protein Interaction Assay

To investigate direct binding between GPNMB and aSyn, we developed a 96-well nickel plate ELISA assay. The Pierce Nickel Coated ELISA plate (ThermoFisher) was first washed five times with 200 uL of wash buffer per well (10 mM Imidazole, 50 mM Phosphate buffer, 300 mM NaCl, 0.05% Tween-20, pH 7.4). Based on the manufacturer’s reported binding capacity per well, 9 pMol of “bait” protein, which was human GPNMB ECD with a his-tag in the C-terminal direction (MW of 70kD), diluted in 100 uL of coating buffer (10 mM Imidazole, 50 mM Phosphate buffer, 300 mM NaCl, pH 7.4) was loaded per well. The GPNMB ECD-his incubated for two hours at room temperature with orbital shaking, followed by five washes. The plate was then blocked with 100 uL per well of 1% bovine serum albumin (Sigma-Aldrich), 2 mM Imidazole (Thermo Scientific), and 0.05% Tween-20 (BioRad) in PBS (Gibco) (pH 7.4) for one hour at room temperature with orbital shaking. Following five additional washes, the “prey” proteins were added in a twofold molar ratio (18 pMol per well) diluted in 100 uL of blocking buffer. Human wild-type monomeric alpha-synuclein and pre-formed fibrils were provided by Dr. Kelvin Luk. The appropriate molar ratio for both forms of alpha-synuclein were calculated using the molecular weight for the monomer (14 kD). The “prey” proteins were incubated for two hours at 37°C with orbital shaking. The plate was then washed five additional times prior to addition of the biotinylated primary antibodies. The biotinylated anti-GPNMB antibody (sc-271415), biotinylated anti-alpha-synuclein antibody (syn211), and biotinylated Mouse IgG1, κ Isotype Control antibody (BD Biosciences) were diluted to a working concentration of 0.001 mg/mL with blocking buffer. 100 uL of diluted antibody was added to each well and incubated overnight at 4°C with orbital shaking. The following day, the plate was washed five additional times and 100 uL of streptavidin-HRP (1:1000, Jackson ImmunoResearch) diluted in blocking buffer were added to each well. Following a 30-minute incubation at room temperature with orbital shaking, the plate was washed five additional times and 1-Step TMB ELISA substrate (100 uL per well) was added and allowed to incubate for 5-20 minutes. The reaction was stopped using 50 uL of 10% phosphoric acid (LabChem) per well and read at 450 nm on the FLUOstar Omega plate reader (BMG LABTECH).

