## Supplemental Materials for "Secreted GPNMB enhances uptake of fibrillar alpha-synuclein in a non-cell-autonomous process that can be blocked by anti-GPNMB antibodies"

Includes:

- **Figure S1**. GPNMB-aSyn interaction by pulldown assay.
- **Figure S2**. Analysis of single-nuclei RNAseq microglia midbrain datasets.
- **Figure S3**. GPNMB expression in PD vs. control brain: neurons and astrocytes.
- **Figure S4.** GPNMB expression in HMC3 and iPSC-derived human microglia.
- **Figure S5**. Validation of iMicroglia and iNeuron identity based on lineage-specific gene marker expression.
- **Figure S6.** Uptake of aSyn fibrils in iNeurons treated with conditioned medium from iMicroglia.
- **Figure S7**. Screening of anti-GPNMB monoclonal antibodies.
- **Figure S8**. Validation of monoclonal antibody specificity against the GPNMB ECD.
- **Figure S9**. Anti-GPNMB mAb-1 blocks development of aSyn pathology in *GPNMB* KO iNeurons treated with GPNMB ECD or with conditioned medium from wild-type iMicroglia.
- **Table S1**. Human neuropathological brain samples information.
- **Table S2**. List of single-cell RNA-sequencing datasets examined for this study.
- **Table S3**. List of top differentially upregulated genes in PD microglia (integrated dataset).
- **Table S4**. List of top differentially upregulated genes in PD microglia (midbrain).
- **Table S5**. Characterization of anti-GPNMB monoclonal antibody activity.
- **Table S6**. Demographic and pathologic characteristics of the genotyped neurodegenerative disease cohort.

Additional supplementary materials not included in this file:

- Cell Profiler pipeline
- Image J macro automated script
- Script for analyses of single-cell RNA sequencing data and manual cell type annotation (on Github)

**Supplementary Figure 1.**

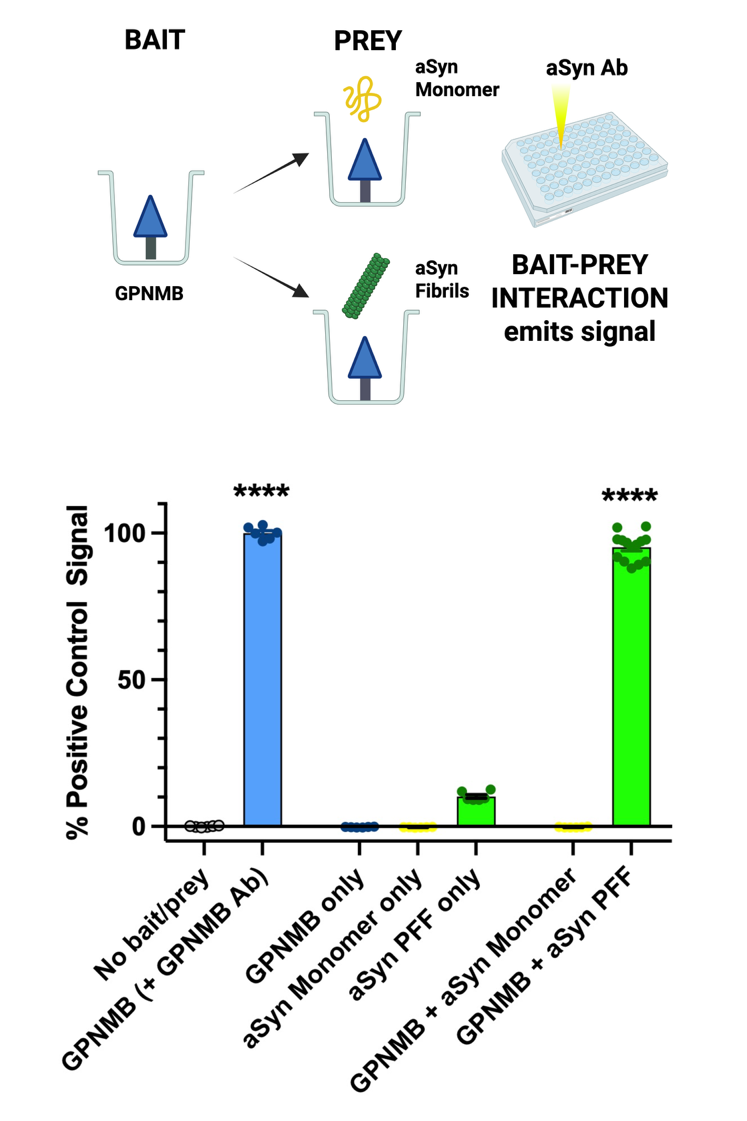

**Supplementary Figure 1. GPNMB-aSyn interaction by pulldown assay.**

**Top:** The GPNMB ECD is used as bait, and alpha-synuclein (aSyn), in monomeric and fibrillar forms, is tested for interaction as prey. The anti-aSyn antibody syn211 (Abcam) recognizes both monomeric and fibrillar forms of captured aSyn and emits signal.

**Bottom:** GPNMB ECD captures aSyn fibrils (GPNMB + aSyn PFF) efficiently, to nearly 100% of the signal detected by anti-GPNMB antibody (compare blue bar to right-most green bar). In contrast, GPNMB ECD does not capture aSyn monomer (GPNMB + aSyn Monomer). Bait alone (GPNMB only) or prey alone (aSyn Monomer only, aSyn PFF only) emits no significant signal when detected with anti-aSyn antibody. Each dot represents one well, with replicate wells distributed across experiments conducted on 3 different days. All data are normalized to the GPNMB bait, detected by the anti-GPNMB antibody sc-271415 condition (Santa Cruz), to account for experiment-to-experiment variability in total signal. ****p<0.0001 nested one-way ANOVA, followed by pairwise comparisons corrected for multiple hypothesis testing.

**Supplementary Figure 2.**

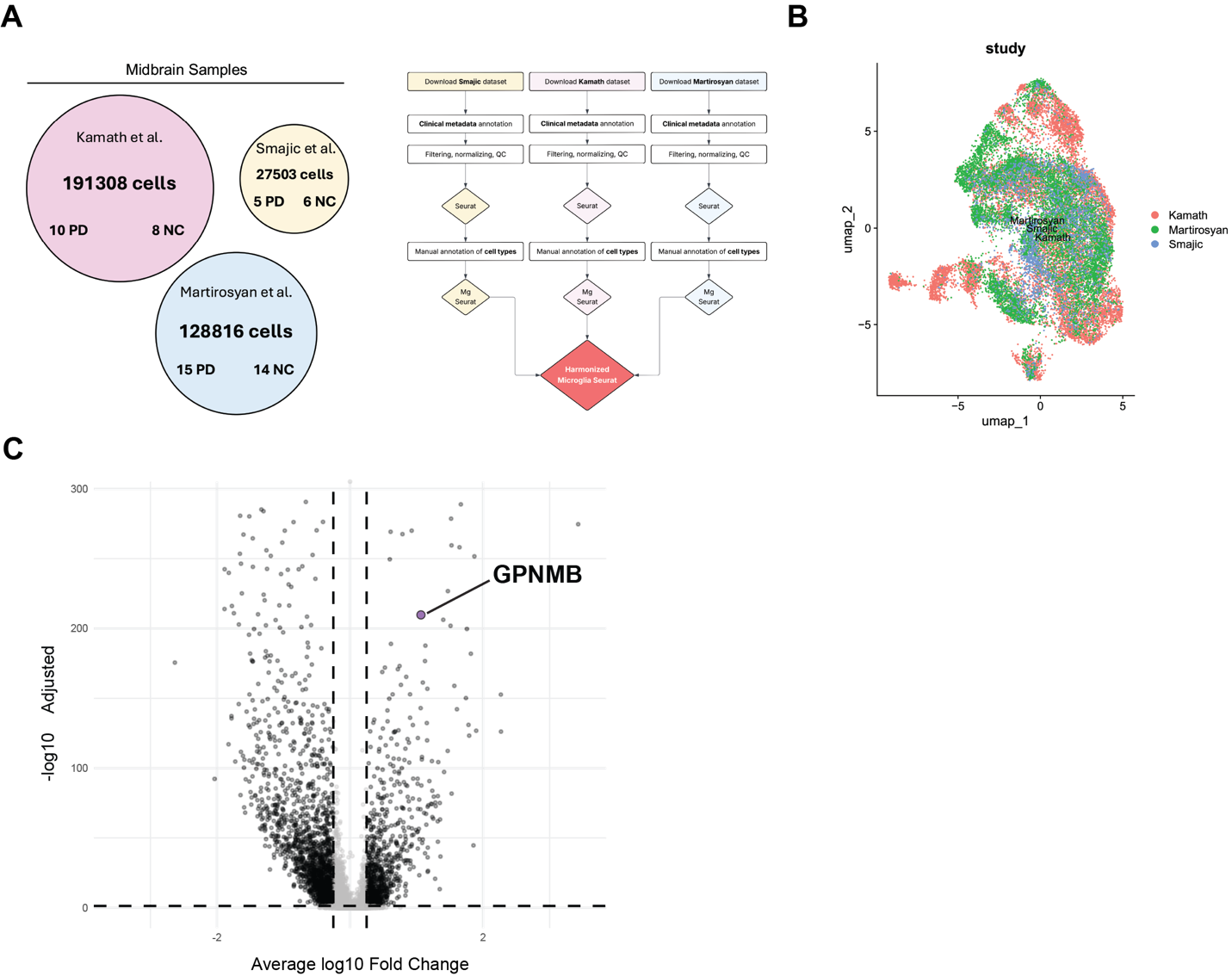

**Supplementary Figure 2. GPNMB is among the top upregulated microglial genes in the midbrain of individuals with PD.**

**(A)**. Pipeline for filtering and integration of microglial gene expression from three midbrain snRNA-seq studies comparing PD individuals with neurotypical controls.

(**B**). Resulting UMAP projection from the integration of the midbrain datasets, which shows reduction of batch effects and overlap between studies.

**(C).** Volcano plot showing the differentially regulated genes comparing PD and NC midbrain microglia. GPNMB is highlighted as one of the top 20 upregulated genes in PD microglia.

**Supplementary Figure 3.**

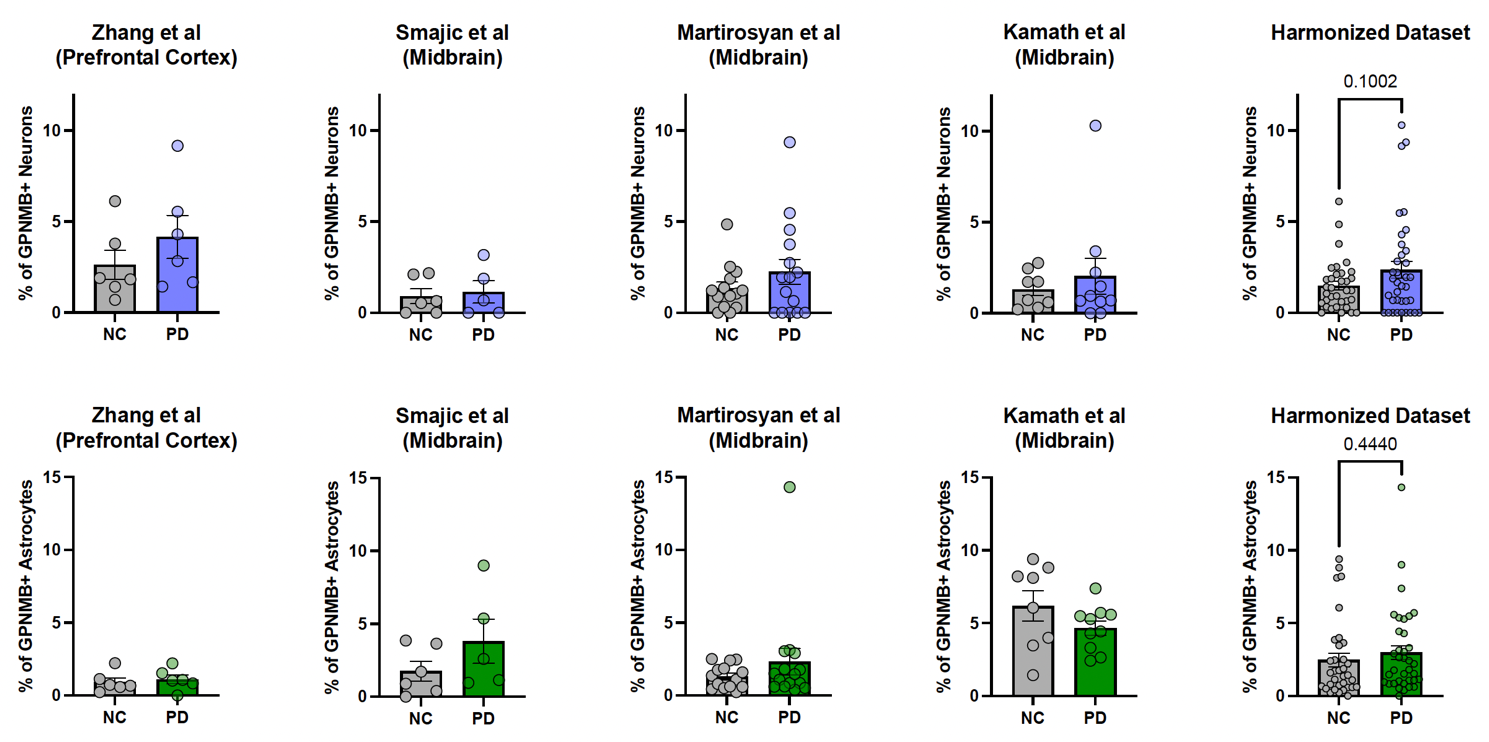

**Supplementary Figure 3**. **GPNMB expression in PD vs. control brain: neurons and astrocytes.**

Percentage of neurons (top) and astrocytes (bottom) expressing detectable levels of GPNMB transcript, stratified by diagnosis (PD or NC) from each scRNAseq study, as well as the integrated data from all four studies. In contrast to microglia, neurons and astrocytes do not differ with respect to the proportions expressing GPNMB in PD vs. NC.

**Supplementary Figure 4.**

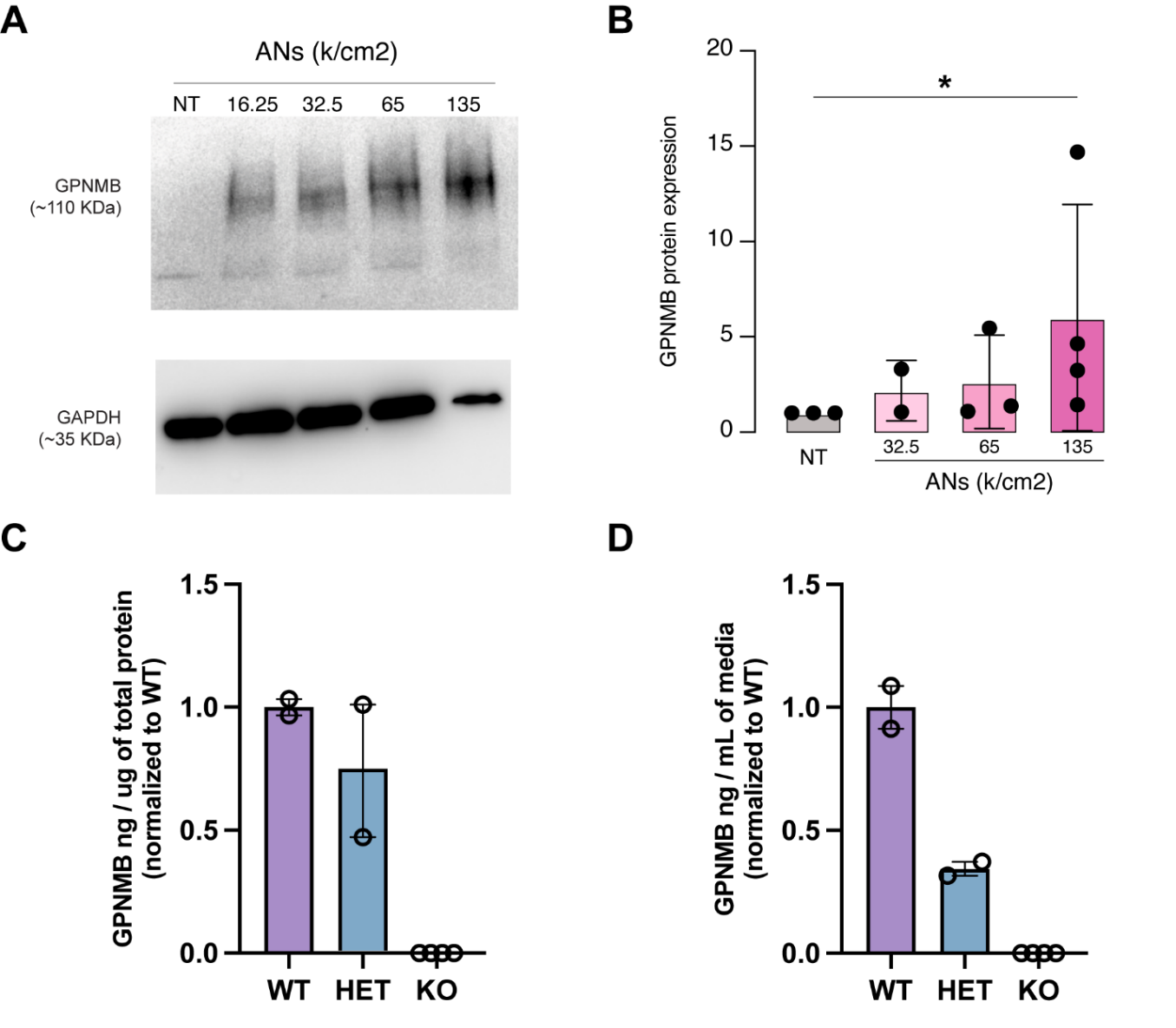

**Supplementary Figure 4**. **GPNMB expression in HMC3 and iPSC-derived human microglia.**

**(A)**. Representative Western blot showing dose-dependent increase in GPNMB protein expression in response to treatment with increasing doses of apoptotic neurons (ANs) compared to non-treated (NT) controls, which have no detectable GPNMB expression. GAPDH is shown as a loading control.

**(B)**. Quantification of GPNMB protein levels normalized to GAPDH as loading control after treatment with different doses of ANs. AN treatment at 135k/cm^2^ significantly increased GPNMB protein expression compared to NT controls (P < 0.05). Data are presented as mean ± SEM, with each dot representing one well. Statistics were calculated using a one-way ANOVA.

(**C and D**). Quantification of GPNMB protein levels in lysate and conditioned media, measured by ELISA, in wild-type (WT), GPNMB heterozygous knock-out (HET) or GPNMB knock-out (KO) iMicroglia.

**Supplementary Figure 5.**

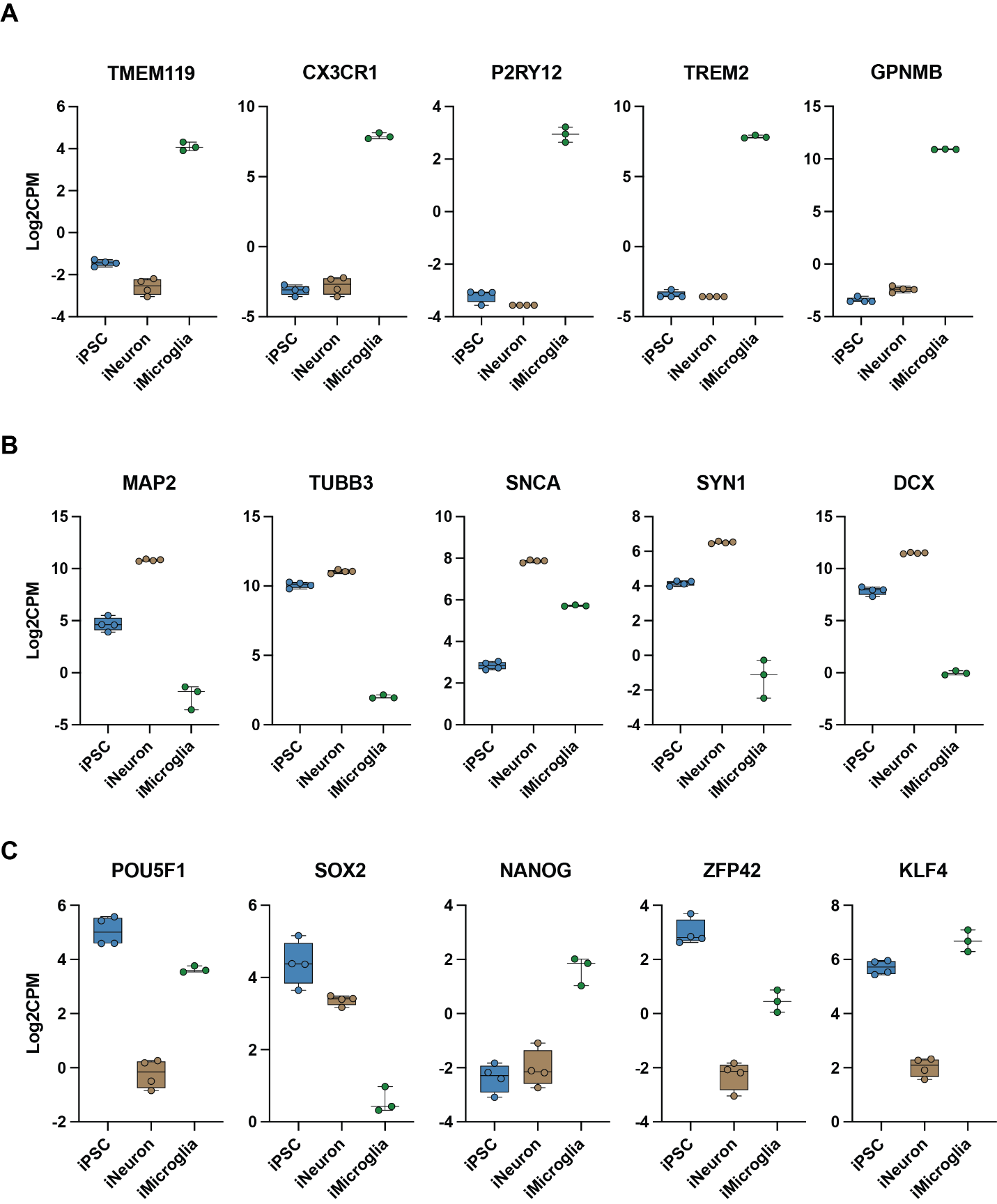

**Supplementary Figure 5**. **Validation of iMicroglia and iNeuron identity based on lineage-specific gene marker expression.**

**(A)**. iMicroglia identity was confirmed by high expression of canonical microglial markers (*TMEM119*, *CX3CR1*, *P2RY12*) and disease-associated markers *TREM2* and *GPNMB*. As expected, *GPNMB* transcript levels are high in iMicroglia compared to iNeurons, with negligible expression levels in iPSCs. Data are presented as log2 counts per million (CPM) for each cell type.

**(B)**. iNeuron identity was confirmed by high expression of neuronal markers (*MAP2*, *TUBB3*, *SNCA*, *SYN1* and *DCX*) compared to iPSCs and iMicroglia. Data are presented as log2 counts per million (CPM) for each cell type.

**(C).** Pluripotency markers (*POU5F1*, *SOX2*, *ZFP42*, *KLF4*) were enriched in undifferentiated iPSCs compared to differentiated iNeurons and iMicroglia. Data are presented as log2 counts per million (CPM) for each cell type.

n=3-4 replicates for each cell type, profiled by RNA-seq.

**Supplementary Figure 6.**

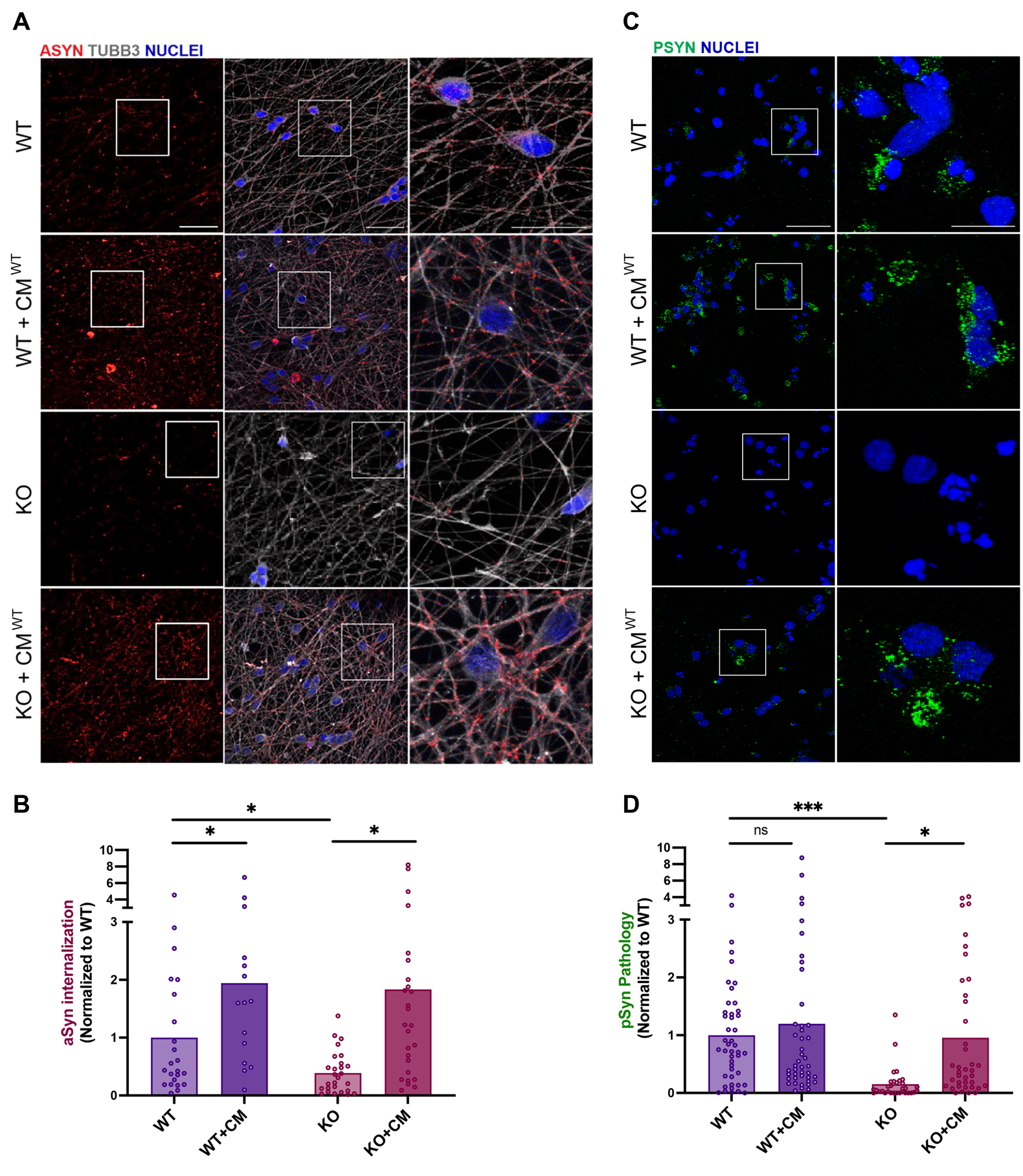

**Supplementary Figure 6. Uptake of aSyn fibrils in iNeurons treated with conditioned medium from iMicroglia.**

**(A)**. Representative images of iPSC-derived neurons (iNeurons) including wild-type (WT), GPNMB knockout (KO), and both WT and KO iNeurons treated with CM collected from WT iMicroglia (WT + CM^WT^, KO + CM^WT^). Labeled aSyn PFFs internalized by cells are shown in red, while tubulin staining (Tub) is shown in grey, and nuclei stained with DRAQ5 in blue. Scale bars: 50 μm (overview), 20 μm (inset).

(**B**). Quantification of aSyn uptake in neurons exposed to CM^WT^ from iMicroglia. At baseline, KO iNeurons show reduced aSyn internalization compared to WT (**p*=0.015). KO iNeurons treated with CM^WT^ increase aSyn internalization significantly (**p*=0.021). WT iNeurons treated with CM^WT^ also increase aSyn internalization (*p*=0.049). Each dot represents one field, with 3-7 fields per replicate (well, n=5) across 3 differentiations, and bar depicts the mean. Statistics were calculated using nested t-test (one-tailed, given expected direction) to account for non-independent fields (see Methods).

**Supplementary Figure 7.**

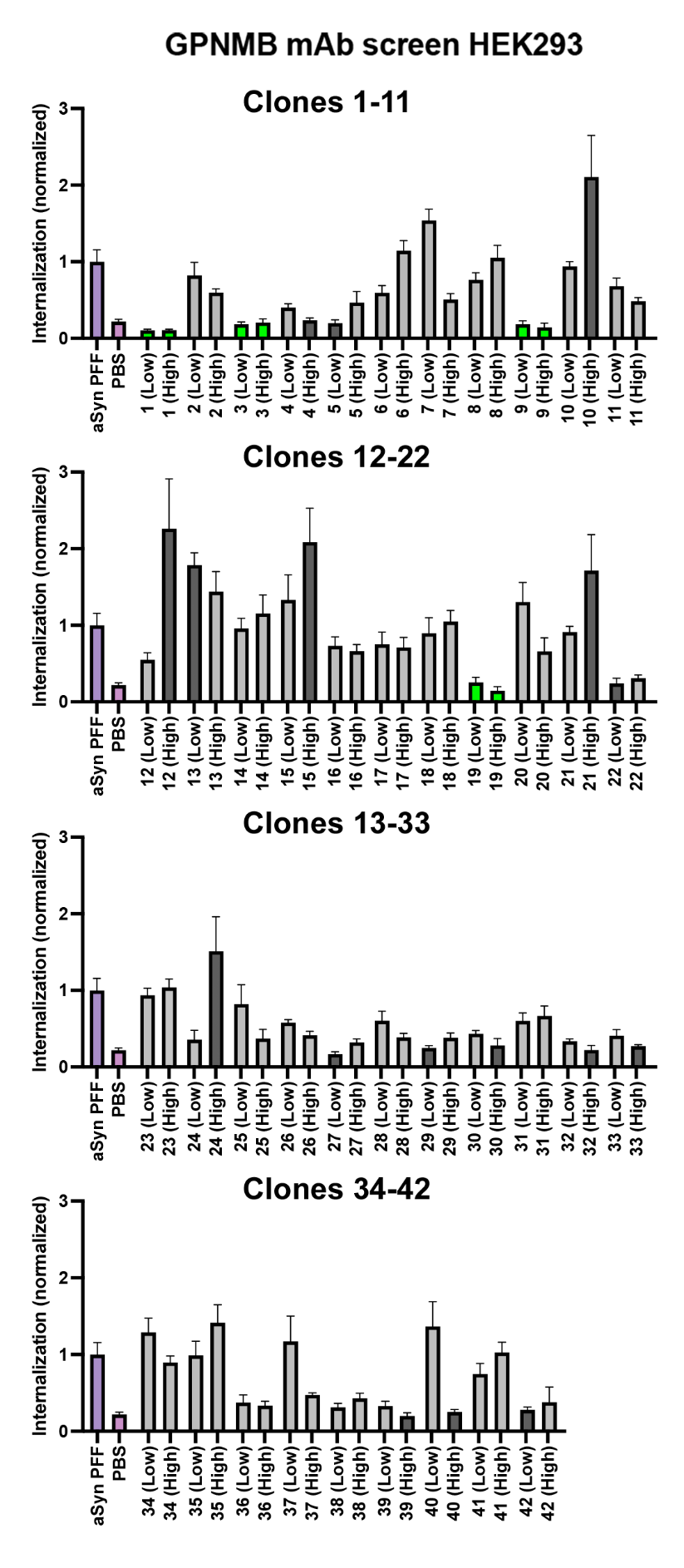

**Supplementary Figure 7**. **Screening of anti-GPNMB monoclonal antibody reveals blockers of alpha-synuclein uptake**.

Quantification of alpha-synuclein fibril (PFF) uptake in HEK293 cells stably expressing GPNMB-GFP following treatment with 42 different monoclonal antibodies (mAbs 1-42) at two different doses (low (75ng/mL): L; high (750ng/mL): H). For each experiment, uptake was normalized to the no-antibody, PFF-treated condition (purple bar). Means +/- SEM are shown. Antibody clones that significantly changed uptake at both doses are colored green, while clones that significantly changed uptake at only one dose are colored dark grey (*FDR-corrected p<0.05). Data were analyzed as a nested ANOVA to account for interdependent (fields within one well) data, with correction for multiple hypothesis testing (FDR<0.05).

**Supplementary Figure 8.**

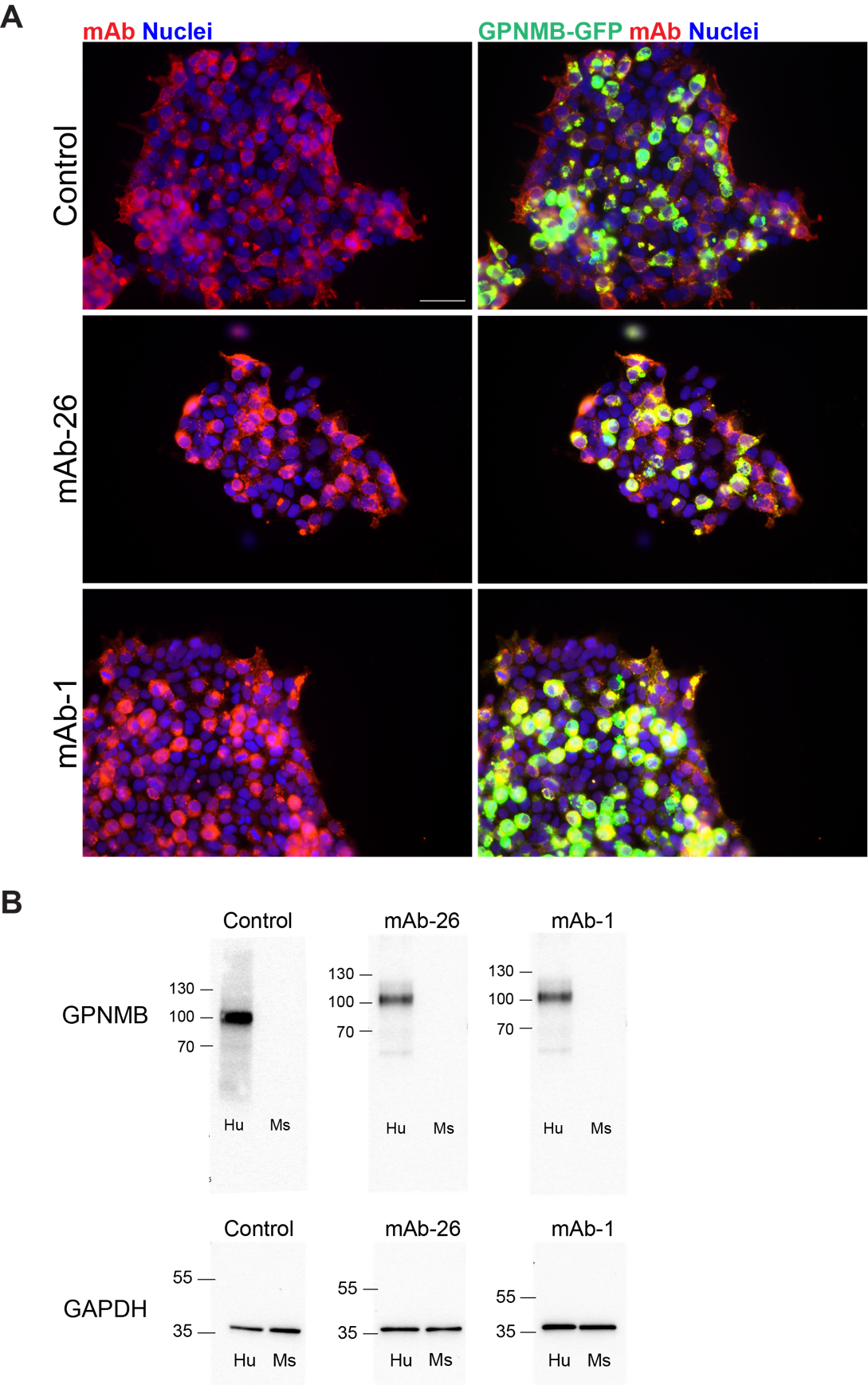

**Supplementary Figure 8**. **Validation of monoclonal antibody specificity against the extracellular domain (ECD) of GPNMB.**

**(A)**. Immunofluorescence images of HEK293 cells stably expressing GPNMB-GFP stained with the commercially available anti-GPNMB monoclonal antibody sc-271415 (Control) or monoclonal antibodies raised against the GPNMB ECD investigated for ability to block uptake of aSyn fibrils or subsequent development of phosphorylated aSyn aggregates (mAb-1, mAb-26). Antibodies targeting the ECD (red) colocalize with the constitutively expressed GPNMB-GFP signal (green). Nuclei are stained with DRAQ5 (blue). Scale bar = 50μm.

**(B)**. Representative Western blot images showing specific detection of GPNMB by the control antibody D9, mAb-1, and mAb-26 in lysates from HEK293 cells over-expressing either human (Hu) or mouse (Ms) GPNMB. Antibodies selectively detected human GPNMB, with no cross-reactivity with the mouse ortholog. GAPDH is shown as a loading control.

**Supplementary Figure 9.**

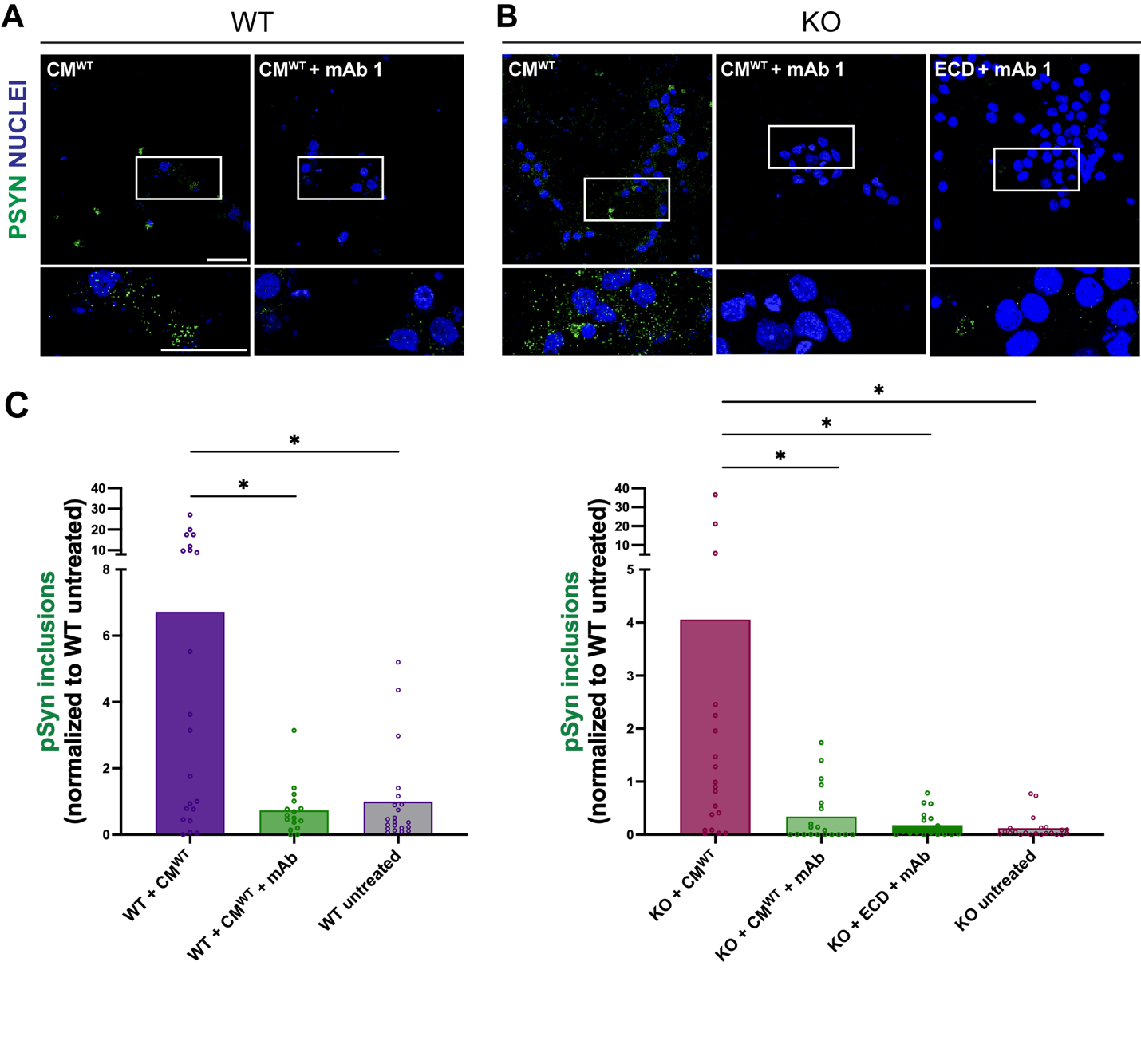

**Supplementary Figure 9. Anti-GPNMB mAb-1 blocks development of aSyn pathology in *GPNMB* KO iNeurons treated with GPNMB ECD or with conditioned medium from wild-type iMicroglia.**

**(A).** Representative images of WT iNeurons treated with either conditioned medium from WT iMicroglia (CM^WT^) alone or in combination with high doses of mAb-1. aSyn pathology aggregates are shown in green, and nuclei are stained with DRAQ5 in blue. Scale bars: 50uM (overview), 20uM (inset).

**(B).** Representative images of KO iNeurons treated with CM^WT^, CM^WT^ in combination with high doses of mAb-1, or GPNMB ECD in combination with high doses of mAb-1. aSyn pathology aggregates are shown in green, and nuclei are stained with DRAQ5 in blue. Scale bars: 50uM (overview), 20uM (inset).

**(C)**. Quantification of mAb-1 effects on development of aSyn pathology in WT (left) or KO (right) iNeurons. In WT iNeurons treated with CM^WT^, addition mAb-1 significantly reduces aSyn pathology (*p=0.018). Addition of mAb-1 similarly reduces aSyn pathology in KO iNeurons treated with either CM^WT^ (*p=0.038) or GPNMB ECD (*p=0.033). For each genetic background (WT, KO), conditions with no CM or mAb-1 treatment are also included as an additional control.

Data for each genetic background were analyzed as a nested one-way ANOVA to account for interdependent (fields within a well) data, followed by pairwise comparisons to discern mAb treatment effects. One-tailed p-values (given expected direction of effect) for indicated pairwise comparisons are presented. Each dot represents one field, with 4-6 fields quantified per replicate (well, n = 4) across 2 differentiations.

**SUPPLEMENTARY TABLES**

**Table S1**. Human neuropathological brain samples used for immunohistochemistry analysis of GPNMB expression across brain regions and diagnoses.

| **Case #** | **Brain region** | **Neuropathological Diagnosis** | **Clinical Diagnosis** | **rs199347** |
| --- | --- | --- | --- | --- |
| 1 | Temporal Cortex | LBD + AD | PDD | GG |
| 2 | Temporal Cortex | LBD | PDD | GA |
| 3 | Temporal Cortex | LBD | PDD | GG |
| 4 | Temporal Cortex | NC | Normal | AA |
| 5 | Temporal Cortex | NC | Normal | GA |
| 6 | Temporal Cortex | LBD | PD | AA |
| 7 | Temporal Cortex | LBD + AD | DLB | AA |
| 8 | Temporal Cortex | LBD + AD | PD | GG |
| 9 | Temporal Cortex | LBD + AD | PDD | GA |
| 10 | Hippocampus | NC | Normal | AA |
| 11 | Hippocampus | LBD | PDD | GA |
| 12 | Hippocampus | NC | Normal | GG |
| 13 | Hippocampus | LBD | PD | GG |
| 14 | Hippocampus | LBD + AD | PDD | GA |
| 15 | Hippocampus | LBD + AD | PDD | AA |
| 16 | Hippocampus | LBD + AD | PDD | AA |
| 17 | Hippocampus | LBD + AD | PD | GG |
| 18 | Hippocampus | NC | Normal | GA |
| 19 | Hippocampus | LBD + AD | PD | GG |
| 20 | Cingulate | LBD + AD | PDD | GG |
| 21 | Cingulate | LBD | PD | AA |
| 22 | Cingulate | LBD | PDD | GA |
| 23 | Cingulate | LBD | PDD | GG |
| 24 | Cingulate | NC | Normal | AA |
| 25 | Cingulate | NC | Normal | GA |
| 26 | Cingulate | LBD + AD | DLB | AA |
| 27 | Cingulate | LBD + AD | DLB | GA |
| 28 | Cerebellum | LBD | PDD | GG |
| 29 | Cerebellum | NC | Normal | AA |
| 30 | Cerebellum | LBD | PDD | AA |
| 31 | Cerebellum | NC | Normal | GA |
| 32 | Cerebellum | LBD | PDD | GA |
| 33 | Cerebellum | LBD | PDD | GA |
| 34 | Cerebellum | LBD + AD | PDD | AA |
| 35 | Cerebellum | NC | Normal | GG |
| 36 | Temporal Cortex | LBD + AD | PDD | GG |
| 37 | Temporal Cortex | NC | Normal | GA |
| 38 | Temporal Cortex | LBD | PDD | AA |
| 39 | Cerebellum | LBD | NA | GG |
| 40 | Hippocampus | LBD | NA | GG |
| 41 | Temporal Cortex | LBD | PDD | GA |
| 42 | Hippocampus | LBD | PD | GG |
| 43 | Cingulate | LBD | NA | GA |
| 44 | Hippocampus | LBD | PDD | AA |
| 45 | Cerebellum | LBD | PD | AA |
| 46 | Cingulate | LBD | PDD | AA |
| 47 | Hippocampus | NC | Normal | AA |
| 48 | Temporal Cortex | NC | Normal | AA |
| 49 | Cerebellum | LBD | PDD | GA |
| 50 | Cingulate | LBD + AD | PDD | GA |
| 51 | Hippocampus | LBD | PDD | GA |
| 52 | Cingulate | LBD | PDD | GA |
| 53 | Cerebellum | LBD + AD | PD | GG |
| 54 | Cerebellum | LBD + AD | DLB | GG |
| 55 | Hippocampus | LBD + AD | PDD | GA |
| 56 | Cerebellum | LBD + AD | PDD | GA |
| 57 | Hippocampus | NC | Normal | AA |
| 58 | Cingulate | LBD + AD | PDD | GG |
| 59 | Cerebellum | LBD + AD | PD | AA |
| 60 | Cingulate | LBD + AD | PDD | AA |
| 61 | Cingulate | NC | Normal | GA |
| 62 | Cingulate | NC | Normal | AA |
| 63 | Hippocampus | LBD | NA | GG |
| 64 | Hippocampus | NC | Normal | CC |
| 65 | Temporal Cortex | LBD + AD | PDD | GA |
| 66 | Temporal Cortex | LBD + AD | DLB | GA |
| 67 | Temporal Cortex | LBD + AD | PD | GA |
| 68 | Temporal Cortex | LBD + AD | PD | GA |
| 69 | Temporal Cortex | LBD + AD | NA | GA |
| 70 | Cerebellum | LBD | NA | GA |
| 71 | Hippocampus | LBD | PDD | GG |
| 72 | Cingulate | LBD | PD | AA |
| 73 | Cingulate | LBD + AD | PDD | GA |

Neuropathological diagnoses include **NC**: Neurotypical Control; **LBD**: Lewy Body Disease; **AD**: Alzheimer’s Disease. Clinical diagnoses include **PDD**: Parkinson’s Disease with Dementia; **PD**: Parkinson’s Disease (no dementia); **DLB**: Dementia with Lewy bodies; **Normal**: Normal Aging. **NA** was used for the cases that had a neuropathological diagnosis without a matched clinical diagnosis. For quantifications used in Figure 2, labels represent neuropathological diagnosis. Some individual cases had multiple brain regions assessed based on availability.

**Table S2**. Publicly available single-cell RNA-sequencing datasets used for cell-specific examination of GPNMB expression.

| **Region** | **Accession #** | **PD (n)** | **NC (n)** | **Total Cells** | **Mg PD** | **Mg NC** |
| --- | --- | --- | --- | --- | --- | --- |
| Midbrain | GSE178265 (Kamath) | 10 | 8 | 191308 | 11132 | 11084 |
| Midbrain | GSE243639 (Martirosyan) | 15 | 14 | 128816 | 9141 | 9694 |
| Midbrain | GSE157783 (Smajic) | 5 | 6 | 27503 | 1099 | 2349 |
| Prefrontal Cortex | GSE202210 (Zhu) | 6 | 6 | 46642 | 3636 | 1617 |

**Table S3**. Top 20 differentially upregulated genes in PD microglia from the integrated dataset including all single-nuclei RNAseq studies (n=4).

| **Rank** | **Gene Name** | **Adjusted p Value** | **Average log2 FC** | **% Expression in PD Mg** | **% Expression in NC Mg** |
| --- | --- | --- | --- | --- | --- |
| 1 | RN7SK | 1.42 E-108 | 1.82 | 11.8 | 6.7 |
| 2 | CHORDC1 | 5.14 E-263 | 1.70 | 32.5 | 20.7 |
| 3 | HSPH1 | 2.655E-218 | 1.69 | 43 | 32.4 |
| 4 | LGALS3 | 6.46 E-173 | 1.66 | 12.2 | 5.5 |
| 5 | PKDCC | 5.88 E-161 | 1.63 | 10.4 | 4.4 |
| 6 | BAG3 | 1.77 E-189 | 1.62 | 15.9 | 8 |
| 7 | HSPA6 | 1.12 E-93 | 1.59 | 11.7 | 6.8 |
| 8 | NUPR1 | 1.30 E-250 | 1.57 | 20.7 | 10.5 |
| 9 | HSPB1 | 6.75 E-236 | 1.56 | 32.6 | 21.5 |
| 10 | CLIC2 | 4.1379E-125 | 1.50 | 13.9 | 7.8 |
| 11 | **GPNMB** | 9.85 E-164 | 1.43 | 23.5 | 15.1 |
| 12 | DNAJB1 | 5.43 E-162 | 1.42 | 21.4 | 13.1 |
| 13 | SERPINE1 | 5.26 E-265 | 1.38 | 19.2 | 9 |
| 14 | PTGES3 | 4.93 E-167 | 1.30 | 48.3 | 39.7 |
| 15 | FKBP4 | 4.98 E-184 | 1.28 | 19.2 | 10.7 |
| 16 | HSPE1 | 4.61 E-37 | 1.23 | 11.9 | 8.9 |
| 17 | AHNAK | 1.05 E-221 | 1.23 | 18 | 8.9 |
| 18 | SNAP23 | 1.83 E-76 | 1.20 | 37.7 | 31.5 |
| 19 | CHI3L1 | 5.00 E-206 | 1.11 | 17.1 | 8.4 |
| 20 | MRPL18 | 1.63 E-76 | 1.10 | 12.4 | 7.8 |

**Table S4**. Top 20 differentially upregulated genes in PD midbrain microglia. For this analysis, the three midbrain-focused single-nuclei RNAseq studies were integrated.

| **Rank** | **Gene Name** | **Adjusted p Value** | **Average log2 FC** | **% Expression in PD Mg** | **% Expression in NC Mg** |
| --- | --- | --- | --- | --- | --- |
| 1 | HSPE1-MOB4 | 3.85E-280 | 3.43 | 8.4 | 1.3 |
| 2 | C15orf26 | 1.26E-131 | 2.27 | 6 | 1.7 |
| 3 | NA.352 | 4.09E-158 | 2.27 | 9.1 | 3.2 |
| 4 | RN7SK | 2.98E-132 | 1.90 | 14.4 | 7.7 |
| 5 | HSPH1 | 3.83E-257 | 1.87 | 43.9 | 32 |
| 6 | BACE2 | 3.97E-50 | 1.86 | 6.1 | 3.2 |
| 7 | LGALS3 | 2.05E-187 | 1.82 | 11.9 | 4.5 |
| 8 | KCNE1 | 1.00E-128 | 1.79 | 7.5 | 2.6 |
| 9 | HSPA6 | 1.93E-136 | 1.78 | 14.1 | 7.2 |
| 10 | BAG3 | 3.90E-205 | 1.76 | 15.9 | 7.1 |
| 11 | PKDCC | 1.18E-155 | 1.74 | 10.9 | 4.4 |
| 12 | HSPB1 | 2.28E-294 | 1.67 | 35.8 | 22 |
| 13 | NUPR1 | 1.27E-263 | 1.65 | 23.2 | 11.4 |
| 14 | CLIC2 | 1.19E-147 | 1.61 | 15.8 | 8.3 |
| 15 | DNAJB1 | 2.12E-164 | 1.57 | 21.6 | 12.7 |
| 16 | AF127936.7 | 3.18E-87 | 1.53 | 7.7 | 3.6 |
| 17 | AHNAK | 5.33E-265 | 1.53 | 16.8 | 6.5 |
| 18 | CHI3L1 | 3.96E-284 | 1.52 | 18.6 | 7.4 |
| 19 | **GPNMB** | 2.01E-207 | 1.51 | 28.1 | 17.2 |
| 20 | FKBP4 | 3.12E-232 | 1.47 | 19.5 | 9.2 |

**Table S5**. Characteristics of the 42 anti-GPNMB mAbs tested.

| **Key** | **Detection Activity** | | **Blocking Activity** | |
| --- | --- | --- | --- | --- |
|  | **ELISA** | **WB** | **Low (Mean ± SEM)** | **High (Mean ± SEM)** |
| **mAb-1** | 1.085 | Yes | 0.1 ± 0.01 | 0.11 ± 0.02 |
| mAb-2 | 1.437 | No | 0.82 ± 0.14 | 0.6 ± 0.06 |
| mAb-3 | 1.94 | Yes | 0.18 ± 0.03 | 0.21 ± 0.04 |
| mAb-4 | 1.723 | Yes | 0.4 ± 0.04 | 0.23 ± 0.04 |
| mAb-5 | 0.746 | No | 0.2 ± 0.05 | 0.47 ± 0.13 |
| mAb-6 | 1.997 | Yes | 0.59 ± 0.1 | 1.14 ± 0.11 |
| mAb-7 | 0.762 | No | 1.54 ± 0.22 | 0.51 ± 0.08 |
| mAb-8 | 0.605 | No | 0.76 ± 0.08 | 3.72 ± 2.64 |
| mAb-9 | 1.171 | No | 0.18 ± 0.04 | 0.12 ± 0.03 |
| mAb-10 | 1.944 | Yes | 0.94 ± 0.16 | 2.11 ± 0.6 |
| mAb-11 | 0.863 | Yes | 0.68 ± 0.16 | 0.48 ± 0.04 |
| mAb-12 | 1.61 | No | 0.55 ± 0.2 | 2.26 ± 0.52 |
| mAb-13 | 1.236 | No | 1.79 ± 0.25 | 1.44 ± 0.23 |
| mAb-14 | 1.753 | Yes | 0.96 ± 0.18 | 1.19 ± 0.27 |
| mAb-15 | 0.868 | No | 1.33 ± 0.24 | 2.09 ± 0.39 |
| mAb-16 | 2.138 | Yes | 0.73 ± 0.12 | 0.66 ± 0.09 |
| mAb-17 | 2.673 | No | 0.75 ± 0.25 | 0.71 ± 0.12 |
| mAb-18 | 0.794 | Yes | 0.9 ± 0.28 | 1.05 ± 0.13 |
| mAb-19 | 1.665 | No | 0.25 ± 0.06 | 0.15 ± 0.05 |
| mAb-20 | 2.651 | Yes | 1.3 ± 0.22 | 0.66 ± 0.21 |
| mAb-21 | 2.08 | Yes | 0.91 ± 0.1 | 1.71 ± 0.47 |
| mAb-22 | 1.173 | Yes | 0.24 ± 0.06 | 0.31 ± 0.05 |
| mAb-23 | 0.935 | No | 0.94 ± 0.1 | 1.04 ± 0.14 |
| mAb-24 | 1.353 | Yes | 0.36 ± 0.12 | 1.69 ± 0.57 |
| mAb-25 | 1.005 | No | 0.82 ± 0.34 | 0.37 ± 0.17 |
| **mAb-26** | 1.927 | Yes | 0.58 ± 0.04 | 0.42 ± 0.05 |
| mAb-27 | 1.27 | Yes | 0.17 ± 0.05 | 0.32 ± 0.03 |
| mAb-28 | 0.731 | Yes | 0.55 ± 0.08 | 0.39 ± 0.06 |
| mAb-29 | 1.128 | Yes | 0.25 ± 0.03 | 0.38 ± 0.07 |
| mAb-30 | 0.803 | No | 0.43 ± 0.03 | 0.28 ± 0.08 |
| mAb-31 | 1.76 | No | 0.6 ± 0.13 | 0.67 ± 0.18 |
| mAb-32 | 0.833 | No | 0.34 ± 0.04 | 0.22 ± 0.05 |
| mAb-33 | 1.274 | Yes | 0.41 ± 0.07 | 0.27 ± 0.05 |
| mAb-34 | 1.254 | No | 1.29 ± 0.22 | 0.9 ± 0.11 |
| mAb-35 | 2.381 | Yes | 0.99 ± 0.2 | 1.41 ± 0.32 |
| mAb-36 | 1.603 | No | 0.37 ± 0.13 | 0.3 ± 0.04 |
| mAb-37 | 2.333 | Yes | 1.17 ± 0.47 | 0.47 ± 0.09 |
| mAb-38 | 1.272 | Yes | 0.31 ± 0.06 | 0.43 ± 0.1 |
| mAb-39 | 2.383 | Yes | 0.33 ± 0.06 | 0.2 ± 0.03 |
| mAb-40 | 1.21 | Yes | 1.37 ± 0.49 | 0.25 ± 0.04 |
| mAb-41 | 0.968 | No | 0.74 ± 0.1 | 1.03 ± 0.14 |
| mAb-42 | 0.84 | No | 0.28 ± 0.04 | 0.38 ± 0.13 |

Relative detection of GPNMB ECD by ELISA, as well as whether each mAb detects GPNMB on immunoblot is listed. Anti-GPNMB monoclonal antibody blocking activity, measured as normalized uptake of alpha-synuclein fibrils in HEK293 cells stably expressing GPNMB-GFP, was quantified by CellProfiler, with means +/- SEM for 10 fields quantified across two replicate wells listed. Blocking activity is normalized to the no-antibody, PFF-treated condition. mAb-1 and mAb-26 (red) are used for downstream experiments in iNeurons.

**Table S6**. Demographic and pathological characteristics of the genotyped neurodegenerative disease cohort. All data is reported as mean (SEM). Proportions are based on primary neuropathological diagnosis.

| **Genotype** | **N** | **% Male** | **Age at death** | **AD (%)** | **LBD (%)** | **CERAD** | **Modified Braak** | **LPCC** |
| --- | --- | --- | --- | --- | --- | --- | --- | --- |
| GG | 313 | 51.11 | 73.30 (0.62) | 43.45 | 14.37 | 1.42 (0.08) | 1.74 (0.06) | 1.00 (0.08) |
| GA | 790 | 57.21 | 73.89 (0.4) | 37.47 | 20.63 | 1.44 (0.05) | 1.74 (0.04) | 1.21 (0.06) |
| AA | 572 | 55.59 | 74.14 (0.48) | 33.91 | 23.77 | 1.42 (0.06) | 1.67 (0.05) | 1.31 (0.07) |
